# Spatial Neurophysiological Gradients at High Resolution Differentiate Tumor Types in the Human Cortex at the Tumor-Brain Interface

**DOI:** 10.64898/2026.10.05.26364665

**Authors:** Thomas Nelson, Brian Hsueh, Peter Hadar, Ipsita Das, Brian F. Coughlin, Ayan S. Mandal, Tommy Wilson, Emory Peng, Andrew Sho Perley, Haziq Rabbani, Roberto Ciordia, Jasmine Zou, Alan Bush, Matteo Vissani, Hazem Ghaith, James Cahill, Jimmy C. Yang, William Muñoz, Erickson Torio, Adam C. Glaser, Joshua D. Bernstock, Kelly L. Collins, Daniel R. Cleary, Harmanpreet K. Tiwana, Alexandra J. Golby, Pamela S. Jones, Bryan D. Choi, Brian V. Nahed, Ahmed M. Raslan, Ziv M. Williams, Shadi A. Dayeh, Pranav Nanda, R. Mark Richardson, Wenya Linda Bi, Steven Tobochnik, Daniel P. Cahill, Gavin P. Dunn, Sydney S. Cash, Angelique C. Paulk

## Abstract

Brain tumors interact bidirectionally with neural circuits, but direct electrophysiological evidence in the human brain across tumor types remains limited. We analyzed intraoperative clinical, mesoscale, and high-density surface electroencephalography recordings from 100 participants undergoing tumor resection, epilepsy surgery, or deep brain stimulation. Population-averaged measures poorly separated diagnoses, whereas spatially resolved aperiodic exponent and high-gamma activity varied with both tumor distance and nearby tumor volume fraction across scales. Gradients differed among IDH-mutant glioma, IDH-wildtype glioblastoma, and metastases. Connectivity further distinguished infiltrative gliomas: beta coherence across tumor–brain connections was reduced in IDH-mutant tumors (p=0.0006; Kruskal–Wallis; mean coherence: tumor–brain 0.26, tumor–tumor 0.44, brain–brain 0.34), unlike IDH-wildtype tumors or metastases. Physiological abnormalities extended into radiographically normal-appearing brain, indicating tumor-type-dependent physiological signatures that may dynamically map the tumor–brain interface complementing structural imaging, with future possibilities as a spatial biomarker of tumor infiltration and network organization in humans.

## Introduction

Brain tumors are increasingly recognized as active participants in neural circuits rather than passive, space occupying lesions. In diffuse gliomas, infiltrating tumor cells engage in bidirectional molecular, cellular, and electrophysiological interactions with surrounding neurons that promote both network hyperexcitability and tumor progression. These interactions, first demonstrated in *in vitro*, *ex vivo*, and non-human animal studies^1–12^, are mediated through extracellular signaling molecules, neurotransmitters, and direct neuron–tumor synaptic connections, creating a feed-forward microenvironment that accelerates tumor infiltration and growth. These reciprocal interactions disrupt normal brain function, frequently producing seizures and other potentially detectable electrophysiological abnormalities ^1–5,9,11,13–16^. This change in brain activity is especially prevalent in diffuse gliomas, the most common malignant primary brain tumor of adults ^7,8,17–22^, though it can occur with metastasized cancers as well ^23,24^. In *in vitro* and rodent studies ^19,25^, brain tumor cells not only form new excitatory connections directly with neurons, but they increase excitatory synaptic connections between nearby neurons, inducing a hyper-connected mesh of tumor and glial cells ^1,2,5,9,11,13,26,27^. Tumor infiltration affects neuron cell types differently. These changes include increased hyperexcitable excitatory neurons and a reduction in inhibitory GABAergic neurons in non-human animal studies and *in vitro* ^26^, where tumor infiltration is potentially slowed by the inhibitory neurotransmitter GABA and application of anti-seizure medications ^11,13,26^. Therefore, one result of tumor infiltration is a shift away from a healthy excitation-inhibition (E/I) balance to a hyperexcitable and more connected network ^28^. The resultant hyperexcitability, concentrated at the so-called “leading edge” or peritumoral region, is associated with higher rates of tumor growth and may lead to epilepsy, which affects 40-80% of glioma patients ^17,27,29–35^.

A shift in the peritumoral E/I balance as well as changes in connectivity should be detectable in high resolution measures of neural activity. Electrophysiologic parameters provide insight into regions of aberrant neural activity across many disciplines of neuroscience. The most widely use measures of hyperexcitability is high gamma power (70-200 Hz) and the beta/gamma ratio (high gamma band power (HGP; 70-200 Hz) to beta band; 15-30 Hz), where the ratio provides within-patient normalization of excitability. Also, commonly used are the E/I slope, interictal discharge (IID) frequency, and the newer aperiodic parameters including aperiodic exponent^36^. The E/I ratio is dynamic measure, where relative hyperexcitability is indicated by flatter E/I slope, lower aperiodic exponent values, higher IID frequency, higher gamma band power, and lower beta/gamma ratio ^36–39^. Increased high gamma activity and a decreased aperiodic exponent are generally interpreted as reflecting greater excitatory, relative to inhibitory, activity.

Several studies have reported alterations in high-gamma activity and the aperiodic exponent in glioma ^1,37,40–42^. However, findings have not been entirely consistent. For example, magnetoencephalography (MEG) studies of patients with glioblastoma have suggested increased inhibitory activity in networks surrounding the tumor, in contrast to findings from direct intraoperative recordings suggesting a shift toward greater excitation ^43,44^. Moreover, the existing literature has largely examined these relationships at relatively coarse spatial scales and has not systematically considered whether different tumor types, which may employ distinct mechanisms of infiltration, produce differing physiological signatures. Addressing these questions requires fine-scaled, direct measurements of highly localized neural activity in and around the tumor, measurements that can be obtained *in vivo* using high-resolution electrodes during planned surgical interventions ^45–59,59,60^.

This emerging understanding of the interplay between tumor and neuronal activity suggests that functional connectivity, the degree to which neural activity is coordinated between two spatially distinct locations, may differ within and outside infiltrating tumors ^61^. Specifically, regions within an infiltrating tumor may exhibit stronger connectivity than regions outside the tumor based on *in vitro* and non-human model studies ^1,2,11,41^. Functional MRI studies have already demonstrated differences in functional connectivity relative to the tumor, though the spatial variation is poorly understood ^62^. Characterizing these spatial patterns could provide a powerful approach for identifying differences in the mechanisms of infiltration across tumor types and other brain lesions ^61^.

Notably, brain tumors are diverse with highly variable prognoses and differing tumor infiltration patterns, likely leading to differences in adjacent neural activity ^6,18,19,63–70^. To contextualize classification, brain tumors are diagnosed using histologic or morphologic features as well as key molecular alterations. Some genetic changes likely serve a critical role in early tumor formation and later growth - for instance, isocitrate dehydrogenase (IDH) mutations – albeit probably to a lesser degree than classical driver mutations of systemic cancers such as anaplastic lymphoma kinase (ALK) for a subset of lung cancer ^24,66,68,71,72^. Furthermore, there is an incomplete understanding of the relationship between these alterations and patterns of neuronal integration; that knowledge gap currently limits diagnostic insight and therapeutic exploitation. For instance, glioblastomas have aggressive infiltration of the tumor into brain tissue and harbor characteristic intra- and intertumoral heterogeneity ^6,25,63,65,70^. On the other hand, gliomas with IDH mutations, though typically slower growing, are more likely to induce epilepsy, with surgical resection and treatment offering high rates of seizure freedom ^12,17,20,29,30,32,33,73^. This slower progression is believed to contribute to more widespread neuronal integration, which may in turn produce more distant hyperexcitability. Additionally, in regions of tumor integration, dysfunctional IDH protein leads to accumulation of 2-hydroxygluterate (2-HG) that putatively acts as an excitatory neurotransmitter. All told, these microscale changes could, and likely do, induce local and network-wide changes in brain activity that could be registered at the scale of high resolution neural devices ^46,47,52,53,74–76^.

We set out to test three primary hypotheses regarding tumor-related changes in brain activity: (1) tumor-associated neural activity exhibits electrophysiological signatures of altered excitatory–inhibitory balance and connectivity that distinguish brain tumors from epilepsy and movement disorder recordings and these physiological signatures further distinguish major tumor classes; (2) these physiological signatures are detectable across recording scales, from conventional clinical ECoG to high-density µECoG; and, most importantly; (3) these signatures vary systematically with spatial proximity to tumor infiltration, becoming most pronounced near the infiltrative tumor margin.

To test these hypotheses, we mapped neural activity to the three-dimensional structure of the brain, tumor infiltration, and other pathology (e.g. edema) using a newly developed modular pipeline (Advanced Localization and Imaging Guidance for Neurosurgical Electrode Recording, ALIGNER) to account for the features of location, lesion type, and recording location ^77^. Metrics of hyperexcitability evaluated here include the aperiodic exponent ^36^; power at different frequency ranges including alpha, beta, and high gamma power; connectivity measured using correlation, coherence, and debiased weighted phase lag index (dwPLI); and detected epileptiform interictal discharges (IIDs). Distinct signatures allowed us to differentiate diagnosis including tumor subtype and even distance to the tumor using machine learning classification tools, particularly differentiating IDH mutant gliomas from GBMs and other diagnoses, indicating passive intraoperative sampling can provide a powerful spatially and temporally high-resolution map of tumor-induced changes in neural activity.

## Results

Human intracranial surface brain recordings (N=101 intraoperative cases) were acquired from 100 participants (mean=50.8 years, range 22 to 80; 50 females, one individual participated in 2 separate cases) undergoing surgical resection of cortical tissue for tumor or epilepsy or implantation of a deep brain stimulation (DBS) device for movement disorders (MD). Patients were defined by groupings that align with World Health Organization 2021 Classification of CNS Tumors ^78^. The first analytical separation was into IDH-mutant (IDHm) versus IDH-wildtype (IDHwt) primary brain tumors, followed by tumor grading. Importantly, grade was determined using integrated pathology that incorporates both histologic and molecular features, which can identify tumors harboring aggressive genetic features on a background of low-grade morphology even in the absence of high-grade histology. Diffuse astrocytomas lacking IDH mutation (IDHwt) were classified as glioblastoma (GBM). Data from individuals undergoing surgery for epilepsy (not tumor resection) and movement disorders were also recorded using these techniques and serve as comparators. The participant distribution of diagnoses includes: non-tumor epilepsy (hereafter labeled epilepsy; N=34), IDHwt (N=26), IDHm (N=23), metastases (N=12), and movement disorders (N=6) (**Fig. 1; Supplementary Table S1-3; Supplementary Fig. S1**).

**Figure 1.**
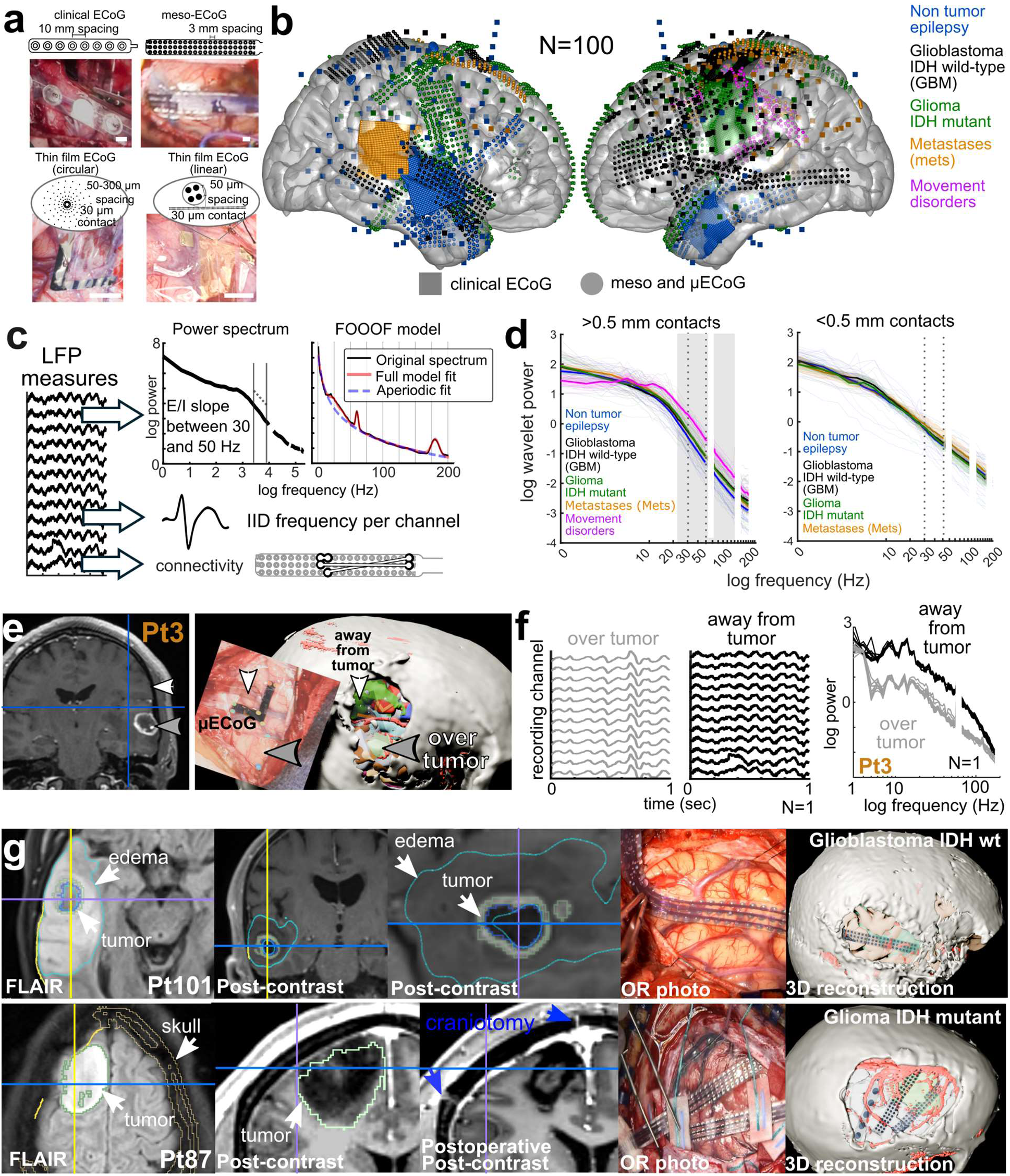
Surface recordings of human brain activity have small differences on the power spectral metrics on the per-participant level without taking into account electrode location. **a.** Schematics and examples of the surface electrocorticography (ECoG) devices used. **b.** MNI-mapped localization of electrodes across cases with color coding as indicated. **c.** Diagram of the neural data pipeline and analytical steps taken. **d.** Power spectral curves calculated using wavelet analyses averaged per-device across channels per-participant (thin lines) and averaged across diagnoses (thick line) with shaded error bars indicating the standard error across the dataset per frequency step, with left from the clinical ECoG (N=64) and meso-ECoG (N=55) and the right curves from the µECoG contacts (N=51). Grey shaded blocks indicate multiple comparisons corrected differences between diagnostic groups per frequency step, p<0.001. **e.** Electrode localization for N=1 participant, Pt3, showing both initial recording site away from tumor and secondary recording site over tumor as identified from a surface visual difference in tissue color and the post-contrast enhancing imaging. **f.** Left and middle: Data from the same participant as in f example of ongoing activity away from the tumor versus over the tumor, N=1, Pt3. Right: Spectral differences across multiple channels (different lines) using the same device for Pt3. **g.** Imaging and photographs used to map electrode locations in two example participants. (top-IDH wt glioblastoma, Pt101; bottom-IDHm glioma, Pt87). GBM-glioblastoma multiforme; IDHm-isocitrate dehydrogenase (IDH) mutation; IDHwt-IDH wild type; mets-metastases; MD-movement disorder.

Surface electrodes used to record brain activity include FDA-approved clinical grids and strips with 3 mm wide contacts and 1 cm spacing (N=61; ECoG), meso-clinical electrodes (N=52; meso-ECoG) that have 1 mm contacts with 3 mm contact to contact spacing ^79^, and thin film µECoG electrodes with 64-1024 channel PEDOT:PSS (poly(3,4-ethylenedioxythiophene) polystyrene sulfonate) or platinum nanorod (PtNR) arrays^45,46,49,51–53,55,60,74,80,81^ (N=48; **Fig. 1**). Some of the physiological recordings used here have been included in previous publications with different types of analyses ^45,46,49,51–53,55,60,74,77,80,81^ (**Supplementary Table S1-3**). This dataset includes a total of 11,086 clinical ECoG, meso-ECoG, and µECoG contacts recorded between 2 to 15 minutes (**Supplementary Table S1; Fig. 1; Supplementary Fig. S1**). Coverage includes mostly frontal and temporal lobe regions and both hemispheres across diagnoses except movement disorder cases that involved a focus on the left frontal lobe (**Fig. 1; Supplementary Fig. S1-5**).

### Power Spectra Are Similar Across Diagnoses Without Spatial Localization

Considering pooled datasets across devices, diagnoses, and surgeon-specific approaches, we first determined if, on the participant and group level, there were commonalities in the neural signal, particularly before considering spatial locations relative to pathologies. When comparing normalized power spectral curves across diagnoses, we found no clear between-tumor differences in spectra (average spectral curves normalized across frequencies from 1 to 200 Hz; see **Methods**; **Fig. 1; Supplementary Fig. S6**). While the power spectra are not obviously different across diagnoses, measures of hyperexcitability were interrogated for per-participant and per-contact differences.

To determine if there are differentiable baseline signatures between IDHm, GBM, metastases, and epilepsy cases on the per-patient level, we averaged the power spectral measures and IID rates across the electrode array per participant. We found there were no significant differences between groups that survived multiple comparisons corrections (threshold with p≤0.0031; Kruskal-Wallis; Bonferroni correction). We further tested using a multi-way ANOVA and again found no significant differences in the binned high gamma, alpha, beta power, aperiodic exponent, E/I slope, beta/gamma ratio, and IID rates with the added confound of general anesthesia versus monitored anesthesia care (MAC) on the level of electrode devices (p=0.21 across comparisons; **Table S1; Supplementary Fig. S7**).

When examining individual electrodes instead of a group average, we do find significant differences between diagnoses for all metrics (**Supplementary Fig. S8**). To understand which of these factors can explain per-electrode variance most consistently, principal component analysis was run on factors standardized using Z-scoring and directional alignment. A set of core components comprising IID frequency, E/I slope, gamma power, and beta/gamma ratio outperformed an expanded set including the core set plus aperiodic exponent, knee, and offset; the first principal component explained approximately 81% of variance using core metrics alone (**Supplementary Fig. S9**). Epilepsy and IDHm glioma cases showed the greatest proportion of hyperexcitability whereas glioblastoma spanned the widest range. Thus, spatially localizing the recording site was necessary to detect major differences between diagnoses.

### Glioma infiltration aligns with spatially localized maps of hyperexcitability

Despite limited differences in power spectral values between participants, we noted a strong spatial gradient in the recordings with respect to radiologic tumor margins for a single example case when moving the electrode to different sites **Fig. 1**; see **Methods**). To quantify and map these spatial differences on the brain, the electrode recordings at each position were mapped spatially in 3D relative to the segmented brain surface, vessels, tumors, edema, lesions, or previous resections using a pipeline utilizing imaging and photographs (Advanced Localization and Imaging Guidance for Neurosurgical Electrode Recording, ALIGNER, see **Methods; Fig. 1, 2; Supplementary Fig. S1-5**)^77^. This manual segmentation was first performed in 3Dslicer informed by the presurgical T1 MPRAGE, post-contrast T1, T2, and FLAIR closest to the time of the surgery. The tumor, lesion, and edema segmentations were then reviewed by experienced clinicians (BH, ST, PH, TN, RC, AM) for each tumor type and scan, which were mapped to MNI space (**Fig. 1; Supplementary Fig. S10**).

We found a spatial pattern of aperiodic exponent values relative to tumor margins, where the aperiodic exponent was lower closer to tumor than at distant locations within individual participants across IDHm, GBM, and metastases cases (Pearson’s correlation ρ>0.26; p<0.001; **Fig. 2**). This aperiodic exponent spatial pattern was found in different electrode types (clinical ECoG, meso-ECoG and µECoG), with different tumor diagnoses (IDHm glioma, IDHwt glioblastoma, and some metastases) and across participants and anesthesia level (monitored anesthesia care, or MAC, or general anesthesia, **Fig. 2**).

**Figure 2.**
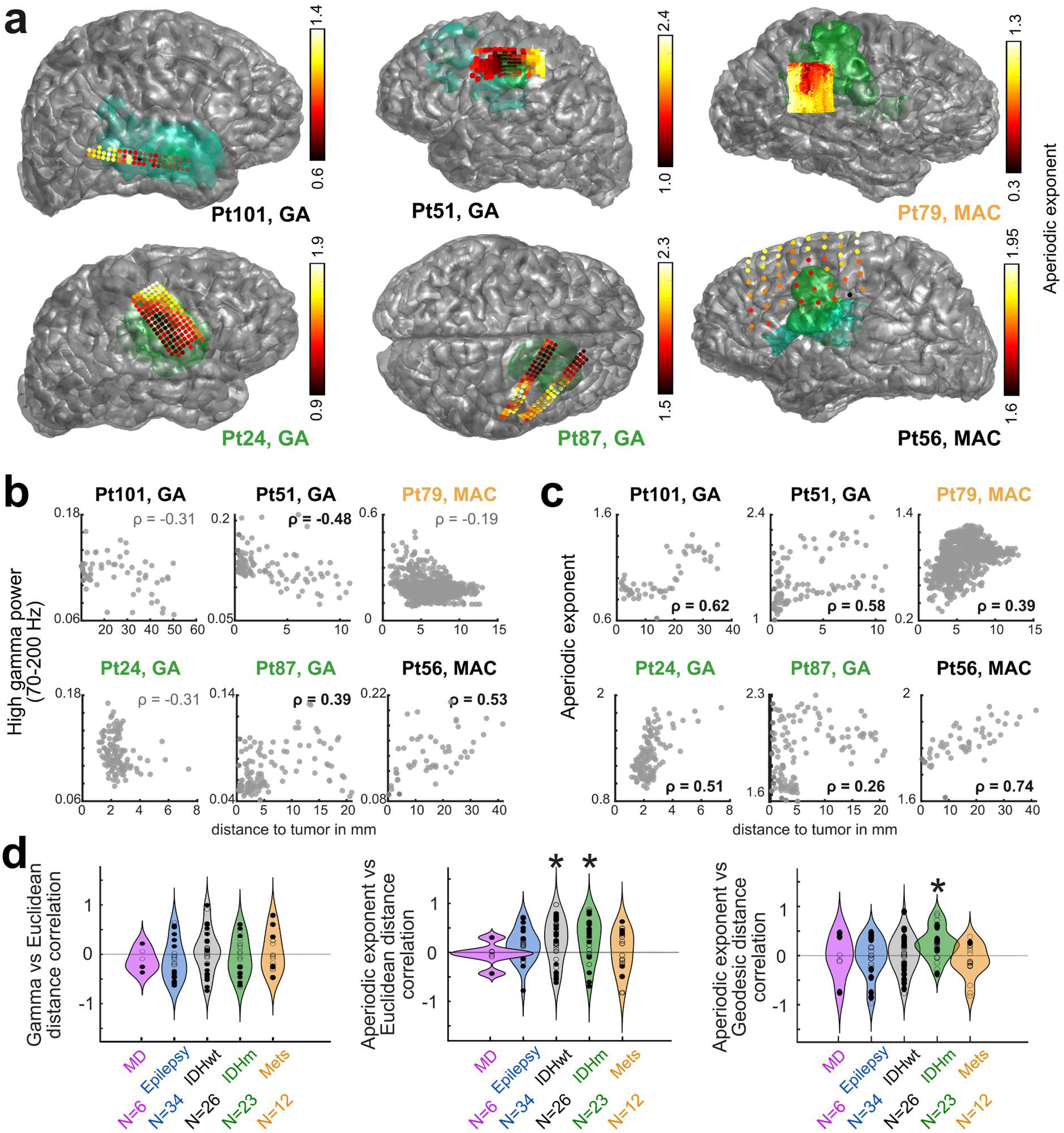
Across participant spatial mapping of spectral features. **a**. Per-participant electrode localizations relative to tumor (green) and edema (teal), with the contacts color coded by measured aperiodic exponent values (color bars in each case). The color coded text per participant (PtXX) indicates diagnoses, namely GBM is black text, green text indicates IDHm, and orange text are metastases cases. **b-c**. Individual examples from the same 6 patients of high gamma (**b**) and aperiodic exponent (**c**) versus Euclidean distance to the tumor edge (top). Rho values are from a Pearson’s ρ correlation with values in bold p<0.001. PtXX indicates a different participant and the type of electrode (e.g. meso-ECoG) is indicated per case. **d**. Across participant correlation values (Pearson’s ρ) between aperiodic exponent and distance from the tumor edge (left) and core (right) for different diagnoses. Dots are per-participant. Asterisks indicate the distribution of correlations are significantly different to zero across participants (p<0.001; Wilcoxon’s signed rank test). Correlation values are between high gamma (top) and aperiodic exponent and Euclidean distance (middle) to tumor edge (left) or core (right) or for the aperiodic exponent correlation with Geodesic distance to the pathology edge or core (bottom). Filled in points are correlation values which are significant (Pearson’s correlation; p<0.001).

To understand spatial patterns across the dataset, to quantify the distances, we calculated both Euclidean and geodesic distances (distances mapped across the cortical surface following the folds of the cortical surface) between electrodes and the pathology (**Fig. 2**; **Supplementary Fig. S11**; see **Methods**)^77^. Assessing both measures allowed us to test the central hypothesis that we should take into account the shapes and curvature of the gyri and sulci (and underlying white matter tracts), which consider the ‘neural’ distance to a tumor, particularly since tumor cells can follow along white matter tracts while electrodes can span lobes ^82–85^ (**Fig. 1,2**). We hypothesized that geodesic distance would have stronger relationships with tumor-induced neural dysregulation. Therefore, we measured both the Euclidean and geodesic distances from the mapped electrode contacts to the edges and centers (core) of the manually outlined pathologies in 3D (tumor, edema, resection, and non-tumor lesion; **Fig. 2; Supplementary Fig. S11**; see **Methods**). For the MD group we created a small 5 mm diameter sphere at the cortical surface location of the inserted DBS lead into the burr hole as a proxy for the lesion (**Supplementary Fig. S11**).

Pooling per-channel data across participants, we did not find strong relationships between distance and the power spectral metrics, both within and across diagnoses, irrespective of whether the distance measures were Euclidean or geodesic or from the edge or the core (**Supplementary Fig. S12-13**). At the per-participant level, though, power spectral metrics, particularly high gamma power and aperiodic exponent, had significant correlations with Euclidean or geodesic distance from the tumor/pathology edge or the core, with a largely positive linear relationship between aperiodic exponent and distance (**Fig. 2**). Importantly, this would imply that aberrant neuronal activity is likely best reflected in relative, rather than absolute, changes over distance from tumor. To investigate this per-participant variation in neural measures, we measured the correlation (Pearson’s ρ) between distance and neural metrics and compared correlations across diagnoses (**Fig. 2**).

We found, overall, per-participant correlations were positive an significantly different to zero for GBM and IDHm gliomas between aperiodic exponent and Euclidean distance to the edge and the core for GBM and IDHm gliomas between aperiodic exponent and Euclidean distance to the edge and the core (p<0.001; Wilcoxon signed-rank test for correlation values across participants significantly different to zero, Bonferroni corrected for multiple comparisons; **Fig. 2; Supplementary Fig. S14-15**). In contrast, we did not find this significant positive trend for epilepsy, metastases and MD cases (p>0.001; Wilcoxon signed-rank test). We further tested the consistency of this trend by evaluating an upper limit of distance for these correlations. We mapped the per-participant correlation between the Euclidean distance to the core vs aperiodic exponent to the minimum distance to the pathology per electrode device. The significant positive correlations were present when electrodes were <50 mm to the pathology, which therefore may represent the detectable boundary using current techniques (**Fig. S16**).

To validate the robustness of this signal to different conditions, we performed a multi-way ANOVA to identify factors that alter this distance versus neural activity correlation; included were channel counts per device, lobe (frontal, temporal, and parietal/occipital), binned minimum distance of the placed electrode to the pathology, seizure history, general anesthesia versus awake status, whether there was a previous resection, and diagnosis. The factors that affected the distance to aperiodic exponent correlation were number of contacts (as there needed to be enough contacts to get a correlation), diagnosis, seizure history, and if the electrodes were within range (<30-50 mm; **Fig. 2**; **Supplementary Fig. S14-16**). Geodesic distance had a lower correlation to aperiodic exponent values (whether to the core or edge) for most diagnoses except for IDHm gliomas. Other neural activity metrics did not show as strong correlations to geodesic distances (core or edge) except for aperiodic exponent and beta/gamma ratio in IDHm gliomas (**Fig. 2; Supplementary Fig. S15**).

In sum, we found power spectral metrics mapped spatially to tumor infiltration on the per-participant level for GBM and IDHm gliomas, particularly relative to the tumor edge and center (core) using Euclidean distances, though IDHm gliomas also showed a strong relationship between the geodesic distance to the tumor edge. These relationships were not as strong for metastatic tumors and could reflect variable or distinct co-option of neural pathways in different types of metastases.

### Physiological Measures Map to Tumor Volume Fraction Around Electrodes

As distance metrics rely on three-dimensional markers to a nearest structure, they may be biased by pathologies with multiple sites of disease, such as brain metastases, or inherently non-uniform shapes like those of primary brain tumors. To estimate the degree of tissue pathology within each electrode’s listening sphere, we calculated Tumor Volume Fraction (TVF) or a non-tumor Lesion Volume Fraction (LVF). We assigned the center points of a matrix of 1 mm width voxels within 5 mm radius sphere around an electrode contact site to brain or lesion (whether tumor, edema, or other pathologies) if the tumor, edema, or other pathology volumes fell within the sphere ^86^ (**Fig. 3**). We then calculated TVF or LVF as the fraction of voxels identified as tumor, edema, or lesion. We also identified the brain volume fraction (BVF) which included the fraction of voxels labelled as within non-pathological brain tissue. Correlating TVF with the aperiodic exponent within individuals (**Fig. 3**), we found significant correlations between the TVF and aperiodic exponent and high gamma power for epilepsy, GBM, and IDHm gliomas, though, the monotonic relationships were not as strong for the beta/gamma ratio, alpha power, beta power, and IID rates for GBMs, epilepsy, and metastases (p<0.001; Spearman’s rho, **Fig. 3; Supplementary Fig. S17-18**).

**Figure 3.**
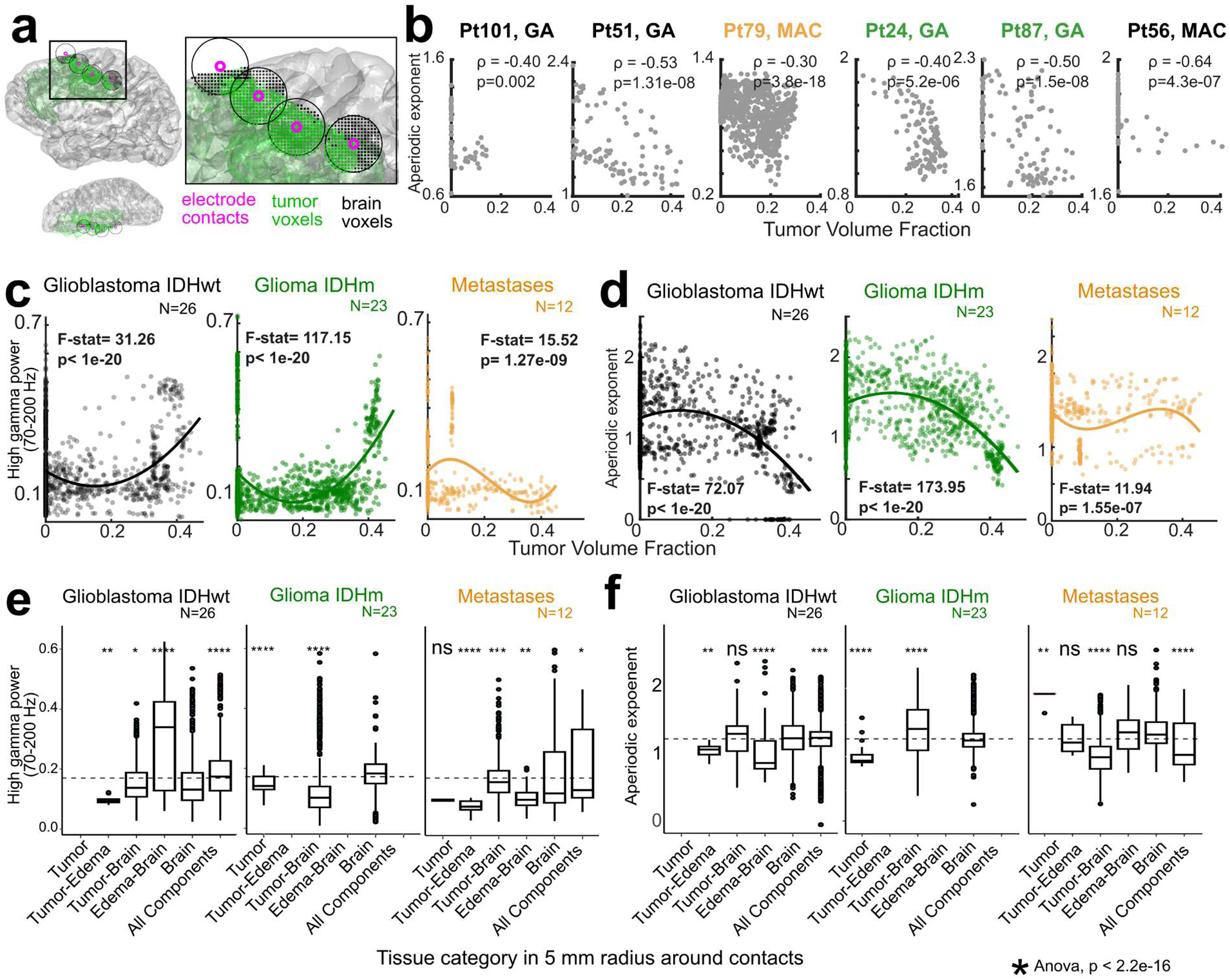
Volume based tissue categorization relationship to neural signals. **a**. Diagram of tumor volume fraction (TVF) or lesion volume fraction (LVF) in a 5 mm radius sphere around each electrode contact, a volume-based categorization of nearby tissue, whether it is tumor (green) or brain volume (black dots). **b**. TVF versus aperiodic exponent for individual recording cases (PtXX designations the same as in Fig. 2). Correlation and p-values: Spearman’s correlation coefficient. The color coded text per participant (PtXX) indicates diagnoses, namely GBM is black text, green text indicates IDHm, and orange text are metastases cases. **c-d**. High gamma power (**c**) and Aperiodic exponent (**d**) versus LVF or TVF around contacts across epilepsy, GBM (IDHwt), and glioma (IDHm) cases. Polynomial fits to the data were calculated with resultant F-statistic results included as well as Spearman’s correlation coefficient (rho) and p-values. N indicates number of participants. **e**. Volume based categorizations relative to physiological measures across IDHm, IDHwt, and metastases (Mets) recordings, with asterisks indicating an ANOVA comparison.

As the relationship also appeared to be nonlinear, we performed a polynomial fit, changing the polynomial line order up to 3 and choosing the order based on the minimum Akaike Information Criterion (AIC) followed by an F-test test to identify the best fit (**Fig. 3; Supplementary Fig. S17-18**). We found LVF versus neural metrics (e.g. aperiodic exponent) was linear for the epilepsy cases while a second or third order polynomial better fit the TVF versus high gamma power, aperiodic exponent, beta and alpha power, and beta/gamma ratio for the IDHm and GBM cases (p< 0.001; F-statistic; **Fig. 3; Supplementary Fig. S17-18**). For metastasis cases, the association with TVF was weaker or not statistically significant for high gamma power, aperiodic exponent, beta and alpha power, IID rates, and beta/gamma ratio (**Fig. 3; Supplementary Fig. S17-18**).

We then categorized the volume around electrode contacts across pathology types within diagnoses. Peritumoral compartments were defined as follows: 1: Tumor only; 2A: Tumor and edema; 2B: Normal-appearing brain and tumor; 3: Edema only; 4: Normal-appearing brain and edema; 5: Normal-appearing brain only; 6: Normal-appearing brain, edema, and tumor (i.e., all components; **Fig. 3**). Use of these categorizations allowed for a richer understanding of regional behavior as well as how hyperexcitability shifts based on the proportions of components. We found that in IDHm tumors, normal-appearing brain has elevated excitatory tone via high gamma band activity and less steep E/I slope; this differs from GBM and metastatic tumors that had generally higher amounts of gamma activity in regions with both brain and edema or with all components (**Fig. 3; Supplementary Fig. S19**). The core metrics were then split into quartiles per tumor group and subtype due to non-normal distributions and increased interpretability. Notably, regions containing normal-appearing brain were richly represented as hyperexcitable, which may implicate more extensive integration than imaging alone would imply (**Fig. 3; Supplementary Fig. S20**).

### Connectivity differentiates glioma subtypes with distance

Considering that tumor infiltration induces hyperexcitable networks in brain tissue^1,2,5,10,11,13,42^, the prediction is that functional connectivity metrics also should differentiate regions relative to the tumor boundary and infiltration. We measured ECoG-based connectivity using correlation, coherence in the theta, alpha, beta, and high gamma bands, and the debiased weighted phase lag index (dwPLI) across contacts at different distances and relative to the various brain regions and pathologies (**Fig. 4, Supplementary Fig. S21-22**). We chose absolute coherence and absolute dwPLI as they fall within the range of 0 to 1 as a measure. Coherence measures both synchronized amplitude of power and the phase relationships between oscillations. On the other hand, dwPLI exclusively measures phase relationships ^87^. Therefore, these metrics could address different mechanisms of synchrony and connectivity.

**Figure 4.**
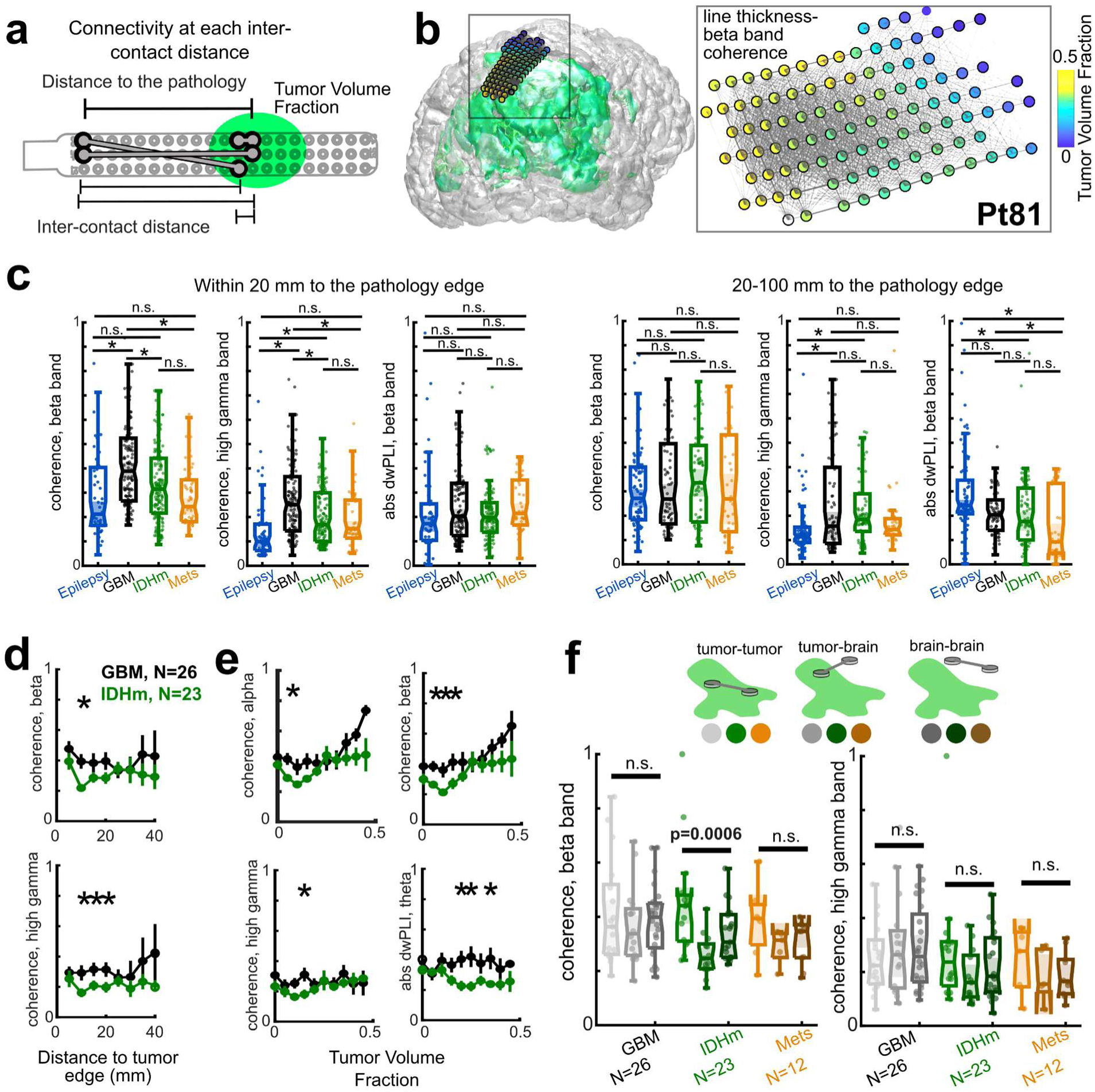
Connectivity differentiates tumor type. **a**. Diagram of connectivity measurements relative to pathology and inter-contact distances. **b**. Example connection strengths between contacts relative to the ratio pathology (tumor) volume on an example brain (IDHwt, left, tumor in green) with box indicating location of the grid (right). Channel contact colors indicate the ratio tumor volume while the thickness of the lines connecting the contacts indicate beta band coherence strength. **c**. Average beta band and high gamma band coherence and beta band absolute weighted phase lag index (dwPLI) dwPLIper patient (dots) and in box plots for the distance to the edge (top) into less than 20 mm (left) and greater than 20 mm (right). p-values indicate pairwise comparisons between groups with p<0.0042, Wilcoxon rank sum test. n.s. indicates p> 0.0042. **d**. The distance to tumor edge binned into 5 mm bins relative to the coherence values for IDHwt and IDHm gliomas. **e**. Binned Tumor Volume Fraction (TVF) relative to the coherence and dwPLI values for IDHwt and IDHm gliomas. For **d** and **e**, p<0.05 significantly different between groups at each binned step and where at least N>5 individuals contributed to each point. Kruskal-Wallis multiple comparisons test. Error bars are standard error. **F**. Boxplots of coherence between pairs of contacts divided into both over tumor (tumor-tumor), one over tumor and one over brain (tumor-brain), and both contacts over brain (brain-brain), color coded per diagnosis as in the legend, with categories based on the Tumor Volume Fraction. Connectivity pairs are averaged per participant (per dot) and compared within diagnoses (GBM, IDHm, and Mets). Statistical comparisons are made between pair categories, Kruskal-Wallis multiple comparisons test.

At the per-participant level, the strength of connectivity measures varied closer to the tumor, with different spatial patterns for coherence and dwPLI (where the distance to the tumor is the average distance of both contacts to the tumor, see **Methods**; **Fig. 4**). Across participants, connectivity between pairs of contacts 5-15 mm apart, which were, on average, 20 mm or less to the tumor or lesion, showed significant differences between diagnoses, with GBMs showing significantly higher alpha, beta, and high gamma coherence, even compared to IDHm gliomas (p<0.0042; Kruskal-Wallis followed by Tukey Pairwise comparisons). Overall, the largest differences in coherence between diagnostic groups were closer to the core or edge of the tumor or lesion (<20 mm). When comparing coherence values, IDHm gliomas were closer to metastases or epilepsy cases (fewer significant differences, also reflected at different inter-contact distances; **Fig. 4; Supplementary Fig. S21-22**). In contrast, dwPLI had larger differences between tumor and non-tumor cases farther from the tumor (>20 mm), with significantly different dwPLI values between tumor and non-tumor diagnoses (p<0.0042; Kruskal-Wallis multiple comparisons, **Fig. 4; Supplementary Fig. S21-22**).

Moving to a higher spatial resolution relative to the tumor, we then examined connectivity at 5 mm binned distances to the pathology core and edge (controlled for inter-contact distance, see **Methods**; **Fig. 4; Supplementary Fig. S23**). Asking whether connectivity varied with this binned distance, we found IDHm coherence was significantly lower than GBMs closer to the tumor, particularly at 10 mm (relative to the edge) and <30 mm (relative to the core). We did not find as striking differences with dwPLI at this scale except for theta band dwPLI. Substituting distance for binned TVF ranging from 0 to 0.5, IDHm alpha and beta coherence values dipped until 0.25 TVF while, above 0.25 TVF, GBM alpha and beta coherence values increased substantially (**Fig. 4; Supplementary Fig. S24**). These results demonstrate a complicated interaction between TVF, distance, and connectivity, which we then examined further.

We subdivided all the network connections into one of three categories: tumor-tumor, tumor-brain, and brain-brain such that contacts with TVF > 0.1 were categorized as tumor and TVF < 0.1 were categorized as brain (**Fig. 4**). Pooling all the connections for these three categories per participant per diagnosis, we found IDHm gliomas had significantly lower theta, alpha, and beta band coherence for the tumor-brain connections than both the brain-brain and the tumor-tumor connections (p=0.0006; Kruskal-Wallis test, followed by Tukey-Kramer pairwise comparisons; **Fig. 4**), but not for high gamma band coherence or dwPLI connections. We did not find differences in between tumor-tumor, tumor-brain, and brain-brain connections for GBMs and metastases. Indeed, a striking result is that tumor-tumor connections in the IDHm, GBM, and metastases cases all had high alpha and beta coherence values while IDHm and metastases tumor-brain and brain-brain connectivity was lower, indicating a spatial difference in connectivity between diagnoses.

### Node strength differentiates diagnoses spatially

To identify if there are specific nodes in the network that have increased normalized connection strength versus other regions, we calculated the node strength (sum connectivity/number of connections) per electrode contact. Node strength varied across the electrode contacts, particularly relative to the tumor in individual cases both in distance to the tumor and TVF (**Fig. 5**). Interestingly, coherence and dwPLI node strength between contacts 5-30 mm apart per frequency range varied inversely in the spatial map. When pooling the data, however, we could recapitulate our distance to connectivity metrics (**Fig. 4, 5**). We found that GBM cases had significantly higher node strength than the IDHm (and Mets) recordings both closer to the tumor and at certain ranges of TVF (**Fig. 5; Supplementary Fig. S25**). Therefore, node strength may be mapped to individual electrodes to classify the distance to tumor along with the power spectral metrics (**Fig. 1–3**).

**Figure 5.**
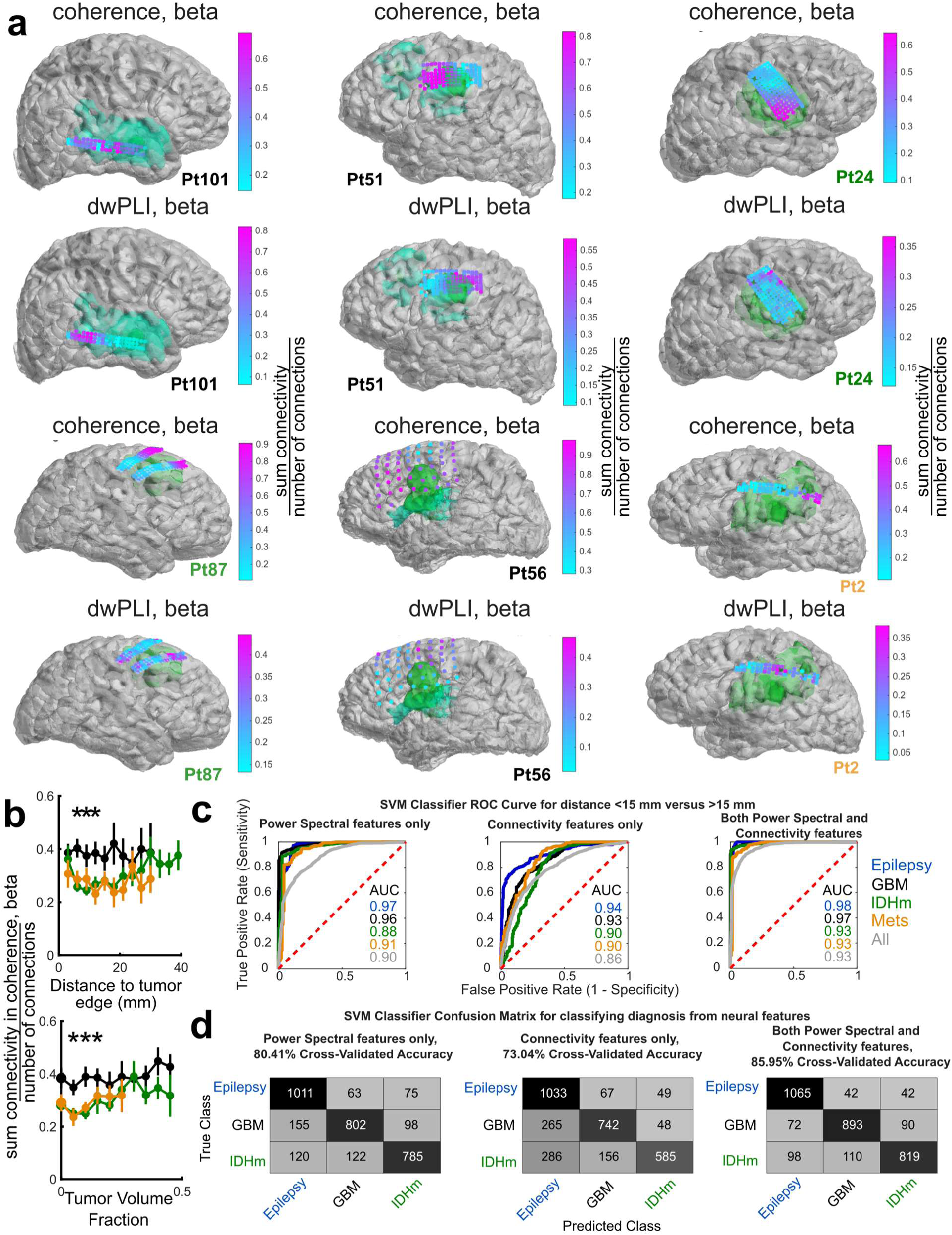
Nodal strength varies spatially and electrode classification is possible for distance and diagnosis. **a**. Per-participant electrode localizations relative to tumor (green) and edema (teal), with the contacts color coded by measured nodal strength per connectivity metric (color bars in each case). The color coded text per participant (PtXX) indicates diagnoses, namely GBM is black text, green text indicates IDHm, and orange text are metastases cases. **b**. Nodal strength connections between contacts between 5 and 30 mm relative to the Euclidean distance to the tumor (top) and Tumor Volume Fraction (bottom). **c**. Support Vector Machine (SVM) model results of predicted versus actual categorizations of nearby (distance <15 mm) and distant (distance to pathology > 15 mm) receiver operating curves (ROC) including power spectral features (left), connectivity features only (middle), and all features (right) subdivided into diagnosis as indicated by the color coding. **d**. Confusion matrices for an SVM model to predict diagnosis from the electrode contacts using power spectral features (left), connectivity features only (middle), and all features (right).

Dividing the distance to the pathology into <15 mm from the edge of the pathology and >15 mm from the edge of the pathology, we explored whether we could classify the contacts within diagnoses and across the epilepsy and tumor diagnoses (**Fig. 5**, see **Methods**). We found the area under the curve (AUC) values were higher for the SVM binary model including the power spectral metrics or the connectivity and power spectral metrics across diagnoses (AUC range of 0.90-0.97 and 0.93-0.98) compared to using only the connectivity metrics (AUC range of 0.86-0.94). Interestingly, we found we could classify electrode contact distances to the tumor or lesion best using power spectral metrics or when combined to connectivity metrics.

We built an SVM multi-class model using an Error-Correcting Output Codes framework to classify the contacts into three different diagnoses, particularly including epilepsy, IDHm, and GBM data (see **Methods**). We found the cross validated accuracy increased with the spectral metrics alone (80.41%) or the combined power spectral and connectivity metrics model (85.95%) versus the connectivity metrics alone (73.04%) (**Fig. 5**). In other words, validation of these findings in subsequent independent cohorts is needed. However, these findings demonstrate that combining metrics may aid in predicting diagnoses.

## Discussion

The emerging field of cancer neuroscience has established that brain tumors interact bidirectionally with the neural circuits they infiltrate: neuronal activity can promote tumor growth, while tumor infiltration can profoundly alter the physiology of surrounding brain ^1–5,9,11,13–16^. Despite these advances, direct electrophysiological evidence of tumor–neural interactions in the human brain remains surprisingly limited ^1,37,40–42,88^. The relatively few studies examining neural activity in and around human brain tumors have reported heterogeneous and sometimes conflicting findings, and it remains unclear whether this variability reflects differences in tumor biology, recording methodology, or the spatial relationship between the recording site and tumor infiltration. Here, we address these possibilities by mapping multiple complementary measures and scales of neural activity to tumor location and infiltration across a large intraoperative electrophysiological dataset. Three principal findings emerge. First, the physiological effects of tumor infiltration are fundamentally spatial: group-averaged electrophysiological measures showed limited diagnostic separation, whereas within-patient changes relative to tumor location were robust. Second, these spatial signatures differed among tumor types, clearly distinguishing between IDH-mutant gliomas, glioblastomas, and metastases likely reflective of differential neural integration. Third, tumor-associated changes were detectable beyond local spectral measures to the organization of functional connectivity, suggesting that infiltration alters not simply the excitability of individual cortical regions but the networks in which they participate. Importantly, there were striking distinctions between IDHm and IDHwt glioma in connectivity, reflecting the differing impact on tumor-brain coherence of these two glioma subtypes. Together, these findings support the concept that tumor infiltration produces a spatially organized and tumor-dependent physiological phenotype that can be measured directly in the human brain.

Perhaps the most important observation is that absolute electrophysiological values alone incompletely capture these effects. Averaging across recording sites or participants largely obscured differences among diagnoses, despite substantial variation among individual contacts. In contrast, relating each contact to the three-dimensional location and extent of pathology revealed reproducible gradients in neural activity. Aperiodic exponent increased with distance from both IDH-mutant gliomas and glioblastomas, consistent with greater relative excitation closer to infiltrated tissue, whereas these relationships were substantially weaker or absent in metastases and the non-tumor comparator groups. Whether the primary site of cancer – e.g., lung versus breast – or tumor genetics may impact the neural integration pattern of metastasis remains unresolved and may dilute any relevant signal in this cohort. Importantly, aperiodic exponent and high-gamma power provided physiological signatures of local tumor burden, varying systematically with the tumor volume fraction (TVF) surrounding each electrode contact, often in a nonlinear manner. These findings suggest that the relevant physiological biomarker may not be an absolute threshold of excitability, but rather the spatial gradient of physiological disruption within an individual brain.

This spatial dependence also provides an important perspective on the concept of the tumor boundary. Many gliomas do not possess a discrete biological edge corresponding precisely to a radiographic boundary; infiltrating tumor cells may extend into tissue that appears structurally normal on conventional imaging ^89–91^. Consistent with this, electrophysiological abnormalities in our recordings were not confined to regions categorized radiographically as tumor. Normal-appearing brain, particularly in IDH-mutant gliomas, frequently exhibited features associated with increased excitatory tone, while GBM and metastatic tumors showed different relationships among tumor, edema, and surrounding brain. Thus, rather than defining a sharp electrophysiological border, these data suggest a physiological transition zone extending across the tumor–brain interface. Whether this transition reflects microscopic tumor infiltration, altered extracellular signaling, synaptic remodeling, network effects propagated from the tumor, or some combination of these mechanisms ^1–5,9,11,13–16,37,40,70^ cannot be determined from the present recordings. Nonetheless, the observation raises the possibility that electrophysiology may provide information about the biological extent of tumor–brain interaction that is complementary to structural imaging. A deeper understanding of this representative extent of tumor involvement could guide future surgical or neuromodulatory strategies. Future work could involve sampling neural activity intraoperatively immediately post-resection to determine if these signals persist.

The differences between IDH-mutant gliomas and glioblastomas are particularly informative. IDH-mutant gliomas showed prominent signatures of hyperexcitability across several measures, consistent with their strong clinical association with seizures and with the prolonged opportunity for slowly infiltrating tumors to integrate with surrounding neural circuits. Yet GBM was not simply a less hyperexcitable version of IDH-mutant glioma. Rather, the two tumor classes exhibited different spatial and network organizations. GBMs demonstrated greater local coherence near tumor, whereas IDH-mutant gliomas showed reduced coherence specifically across tumor–brain connections relative to tumor–tumor and brain–brain connections. These observations suggest that molecularly distinct tumors may reorganize surrounding neural circuits through different mechanisms. Further, connectivity in the beta frequency band reflected key differences in diagnoses, a frequency range which has increasingly been identified as a useful spatial tool in tumor mapping ^40,88^. The resulting physiological phenotype therefore may reflect not simply the amount of tumor present, but the manner in which a particular tumor interacts with and becomes incorporated into the surrounding neural network, particularly in relation to the underlying white matter networks where primary glial tumors develop ^70,82,84,85,92^.

This distinction becomes especially apparent when considering functional connectivity. Much of the cancer neuroscience literature has focused on local neuronal hyperexcitability, but tumor infiltration is hypothesized to alter communication between spatially separated neuronal populations as observed in fMRI and in relation to the white matter network ^28,62,92^. Our findings support this prediction. Coherence and phase-based connectivity measures varied with distance from tumor, TVF, and diagnosis, and the strongest diagnostic differences occurred at particular spatial scales. Notably, tumor–tumor connections showed relatively high coherence across tumor classes, whereas the relationship between tumor and surrounding brain differed markedly by diagnosis. These findings suggest that network topology itself may constitute a physiological signature of tumor biology. Therefore, spectral measures—particularly when combined with connectivity—were highly effective for distinguishing contacts near versus distant from pathology. Local physiology and network organization appear to provide complementary information: the former may be particularly informative about *where* a contact lies relative to structural pathology (lesion or tumor), while the latter may help identify *what kind* of pathological process is present.

An important practical finding is that these relationships were detectable across conventional clinical ECoG, mesoscale electrodes, and high-density µECoG. The spatial pattern of aperiodic activity, for example, was observed across electrode types despite substantial differences in contact size and spacing. This suggests that at least some tumor-associated physiological changes occur at scales sufficiently broad to be captured using clinically available electrodes, while high-density arrays provide the spatial resolution needed to characterize the fine structure of these transitions. Rather than identifying a single optimal recording scale, these findings argue for a multiscale approach: conventional ECoG can provide broad spatial coverage, whereas µECoG and emerging cellular-resolution technologies can interrogate the microscale organization underlying macroscopically detectable abnormalities ^54,75,93^.

These observations also have potential translational implications. Contemporary intraoperative tumor mapping is primarily anatomical and functional: imaging identifies suspected tumor extent, while stimulation mapping identifies cortex that must be preserved ^89,91,94–100^. Electrophysiology could provide a third source of information—a dynamic measure of how the surrounding brain has been altered by tumor infiltration. The ability of electrophysiological features to distinguish contacts near versus distant from pathology provides an initial proof of principle for such an approach. Importantly, this should not yet be interpreted as demonstrating an electrophysiological method for identifying histological tumor margins. Rather, it establishes the possibility that physiological information, integrated with imaging and ultimately with spatially matched tissue pathology, could contribute to a multimodal definition of the infiltrative boundary. The ALIGNER framework provides a means of performing this integration by placing neural recordings, imaging-defined pathology, and surgical anatomy within the same three-dimensional coordinate system ^77^.

Several limitations are important to consider. These recordings were obtained opportunistically during clinically indicated procedures and consequently vary in anatomical coverage, electrode type, anesthesia, tumor location, prior surgery, seizure history, and the number and spatial distribution of contacts. Although the large and heterogeneous dataset allows assessment of the robustness of physiological relationships across real-world surgical conditions, it also limits direct comparisons among diagnostic groups. Radiographic tumor boundaries are themselves imperfect surrogates for microscopic infiltration, making histologically validated spatial sampling an essential next step ^37,40^. The current analyses also identify associations between tumor location and neural physiology but cannot determine whether altered neuronal activity is caused by tumor infiltration, contributes to tumor progression, or reflects reciprocal interactions between the two. Finally, the heterogeneity within even molecularly defined tumor classes makes it unlikely that a single electrophysiological feature will provide a universal biomarker. Prospective studies combining standardized recordings with spatially matched tissue sampling, molecular characterization, and longitudinal outcomes will be required to determine which physiological signatures most directly reflect tumor infiltration and biology ^37,40^.

Together, these findings provide direct evidence that brain tumors are associated with spatially organized alterations in neural physiology in the intact human brain. These changes are not adequately described by a single measure of hyperexcitability or by population-averaged electrophysiology. Instead, tumor-associated pathophysiology emerges from the relationship among local excitability, network connectivity, spatial proximity to tumor, and tumor molecular identity. Mapping these relationships at progressively finer spatial scales - and linking them directly to the molecular and cellular composition of sampled tissue - could provide a bridge between the mechanistic insights of cancer neuroscience and a physiological map of tumor– brain interactions in individual patients.

## Methods

### Human Participants

Human recordings were acquired with 100 participants (mean=50.8 years, range 22 to 80; 50 females; all but 5 individuals right handed **Supplementary Table S1-2**) who were scheduled for surgical resection of cortical tissue as a result of tumor or epilepsy at Massachusetts General Hospital (MGH), Brigham and Women’s Hospital (BWH), and Oregon Health & Science University (OHSU). Most of the patients were already scheduled for a craniotomy for concurrent clinical intraoperative neurophysiological monitoring or testing for mapping motor, language, and sensory regions and removal of tissue (N=89) ^46,46,52,53,77,101^. A subset (N=6) were scheduled for deep brain stimulation (DBS) implantation for movement disorders ^36,79^. Several datasets have been included in previous publications^36,46,46,52,53,77,79,101^.

### Ethics statement

All patients voluntarily participated after fully informed consent according to NIH guidelines as monitored by per-site Institutional Review Boards (IRBs). Massachusetts General Brigham Institutional Review Board (formerly Partners; IRB) provides coverage for Massachusetts General Hospital (MGH) and Brigham and Women’s Hospital (BWH). The Oregon Health Sciences University IRB provides coverage for the Oregon Health Sciences University (OHSU). Participants were informed that participation in the experiment would not alter their clinical treatment in any way, and that they could withdraw at any time without jeopardizing their clinical care.

### Electrodes

When used, intracranial intraoperative clinical electrocorticography were performed simultaneously using PMT or Ad-tech electrodes (Ad-tech Medical, Racine, WI, USA, or PMT, Chanhassen, MN, USA). Ad-tech clinical strip platinum electrodes utilized 10 mm spacing and 2.3 mm contact diameter. PMT Cortac clinical strip platinum electrodes used 10 mm spacing, 3 mm contact diameter. The neurosurgeon placed clinical electrodes in the regions of clinical interest. Clinical electrode recordings used the Natus Quantum system (Natus Neurology Inc., Middleton, WI, USA) or the g.Tech recording system (g.tec medical engineering GmbH Schiedlberg, Austria); the clinical team dictated sampling rates, which were typically at either 4096, 1024, or 512 Hz. When used, intracranial intraoperative clinical ECoG were performed using 64 channel electrodes with 1 cm spacing (Ad-tech Medical, Racine, WI, USA, or PMT, Chanhassen, MN, USA).

Some higher resolution electrodes (meso-grid ECoG) have 3 mm contact to contact spacing, 1 mm wide contact electrodes with either 64 or 128 channels (PMT Corp.) are FDA-approved for use in the operating room and have been used to record neural activity in DBS cases ^79^ and multiple open craniotomy cases. In the DBS cases, the 64 channel PMT grid was slid subdurally from the burr hole created for DBS lead implants on the left side.

Thin film electrode grids were PEDOT:PSS and PtNR grids with specifications detailed in previous publications.^46,52,60,93^ In summary, PEDOT:PSS grids are composed of thin-film poly(3,4-ethylenedioxythiophene) with polystyrene sulfonate ^45,50^. PtNR grids are composed of elemental platinum, the most widely used biocompatible electrode material, and offer greater stability in vivo.^74,102^. Grids measuring 32 × 32 mm with 1 mm spacing (32 x 32 channel square map) or 16 x 64 channels with a 400 mm intercontact spacing were composed of 1024 channels. Grids with 128 channels had 3 conformations: 1) two 64-channel columns with 50-μm pitch; 2) a 4-mm-diameter circular grid.^52,53^

### Data Acquisition

Human intracranial recordings in the OR were made using Natus Quantum System (Natus, Medical Incorporated, Middleton, WI, USA), the Neural Signal Processor (Blackrock Microsystems, Salt Lake City, USA), custom Intan recording system or an Intan 1024 RHD system with a sampling rate of either 20 kHz or 30 kHz (filtered from 1 Hz to 7500 Hz to reduce aliasing; Intan Corporation, Los Angeles, CA, USA). During acquisition, recordings were referenced to sterile ground and reference electrodes (Medtronic or Natus or Rhymlink) placed in nearby muscle tissue or scalp as deemed safe by the neurosurgeon. After the electrode array was laid on tissue and the grounds placed, we recorded ongoing activity. In some cases, a gauze soaked in saline was also placed on top of electrodes for stabilization during the 5-10 minute recordings. For the DBS cases, the reference was an electrode on the other side of the array, facing the dura ^36,79^.

After the electrode array was laid on tissue and the grounds placed, for the thin film recordings, we tested impedances using the Intan system using the RHD2000 series software (Intan Corp.). We used either OpenEphys ^103^ or the RHD2000 software to acquire data on Intan or the Central software when acquiring data with the Neural Signal Processor (Blackrock Microsystems, USA). Trigger signals produced by a custom MATLAB or Python program indicating the hour, minute, and second were additionally sent to both the analog input for the Intan system as well as to the TRIG input to the clinical Quantum system (Natus) which was acquiring the clinical electrode recordings. The clinical Quantum or g.Tec system recorded neural activity at 256, 512, 1024, 2048, or 4096 Hz depending on the intraoperative team choice.

### Intraoperative Video monitoring

Following the patient’s informed consent, we recorded video of the surgery using the overhead camera or took pictures of the craniotomy. The video recordings were acquired by a camera set in the operating room light (Black Diamond Video) or pictures of the preparation were also taken to capture the electrode location.

### 3D Electrode Localization

We developed a pipeline for mapping of electrode location, tumor location, and stimulation location ^77^. Open-source, powerful tools incorporate mapping the MRIs, tumor and brain region localization using segmentation including deep learning, 3D rendering relative to brain surface topology using computer vision, and cloth physics engines within 3D rendering software ^46,52,77,94,95,98,104–115^. This combination of approaches is an innovative step forward pairing imaging, anatomy, function, and future pathological and molecular mapping to neural activity.

Following the surgery, the preoperative T1-weighted MRI was reconstructed using FreeSurfer scripts ^110,116,117^ (http://surfer.nmr.mgh.harvard.edu). The reconstructions were then aligned to images during surgery using blender software (https://www.blender.org/) ^52,118^. The reconstruction and localization of electrodes relative to tumors will involve a series of steps. First, we used either LeadDBS or 3Dslicer (SlicerANTS ^119^) to coregister the multiple MRI scans used to identify tumor location, including the T1-weighted, T1-weighted after gadolinium injection (Gad, with contrast), T2-weighted, and FLAIR MRI scans^105^. We then used Freesurfer, SynthSR, or SynthSeg to perform automatic skull-stripping, cortical and subcortical localization per volume and reconstructed as surfaces to be imported into Blender ^77,104,108–111,118,120–125^. Second, we mapped the tumor extent and boundary through manual segmentation using 3DSlicer^119,126^, visually confirmed by a trained experienced clinicians (BH, ST, PH, TN, RC, AM).

We then use the gyri and sulci, and vasculature (from post-contrast T1 scans) as landmarks with postoperative scans to identify the electrode locations relative to the craniotomy and visible brain structures ^46,47,52,77^ (**Supplementary Fig. S1**). Using to-scale 3D models of the thin film, clinical grids, and Neuropixels arrays (**Supplementary Fig. S1**), we used 3D rendering and modelling tools including geometry nodes and cloth physics to ‘lay’ on the model into the brain in 3D in Blender ^77,104^ as opposed to projection mapping ^127^. We used this approach to create a realistic model with a side by side camera ‘view’ of the electrode location relative to brain regions to optimize positions. Since electrodes were sometimes moved during the recording, the spatial location at each recording time point was also mapped. The electrode recordings were then mapped spatially in 3D relative to the brain surface, vessels, tumors, edema, lesions, or previous resections. Both Euclidean (in MATLAB) and geodesic distances (mapped across the cortical surface, Python using PyMeshLab) were calculated between electrode contacts and relative to the listed pathologies (**Supplementary Fig. S1**). These analyses resulted in arrays of quantified data which included the electrode channel number (per electrode device), the RAS coordinates in the participants’ native space, the distances between contacts and to the pathologies (e.g. tumor or resection) calculated using both Euclidean and Geodesic distance estimations, all in the iEEG-BIDS format imaging combined with electrode and channel information ^77,128^ (https://github.com/Center-For-Neurotechnology/BrainInterface3D).

### Data Analysis

We performed data analysis on the recordings using custom MATLAB (MathWorks, Inc.) or Python scripts. Raw voltage signals were taken directly from the RHD2000 software, OpenEphys, Natus, or Blackrock recordings. We examined recordings and channels for potential noise, produced by the intraoperative environment, that would alter analyses. Channels with excessive line noise, artifacts, or high impedances (>100 kΩ) were removed from analysis ^52^. We then examined the data in 30 second epochs for baseline activity and examined recordings and channels for potential noise, produced by the intraoperative environment, that would alter analyses. Before conducting analysis, LFP sampled above 1000 Hz was decimated to 1000 Hz (lower sampling rates were not changed) and demeaned relative to the entire recording. All the data was common average re-referenced (CAR) per electrode device (e.g. the good channels from the 64 channel PMT grid were averaged and then subtracted from the signal per channel) to remove volume conduction signal and noise overall. Epileptiform activity is detected automatically and confirmed by visual confirmation by experts in the field, including detection of inter-ictal discharges (IIDs)) using automatic packages, programmed in MATLAB (MathWorks, Inc.) or Python ^53,129^.

To remove the effect of 60 Hz noise (and associated harmonics), the power and coherence around each of the noise peaks was changed to not-a-number (NaN) values for 60 Hz (55-65 Hz band), 120 Hz (115-125 Hz band), and 180 Hz (175-185 Hz band) in MATLAB. For the FOOOF analysis (specparam, see below), the 60 Hz signal was not altered at it was already taken into account in the modelling of the spectra for the final calculations ^36^. All the data was then converted into the iEEG BIDS format for easier data processing ^128^.

Spectral curves were calculated using both the Morlet wavelet transform and the Multitaper method at a 1-Hz spectral resolution from 0 to 200 Hz using the Fieldtrip MATLAB package to produce curves which can be used to directly compare with current literature focusing on excitatory vs inhibitory estimates^43,44,130,130–133^. These curves were then averaged across epochs per channel. For the different frequency ranges, the average power for the alpha (8-15 Hz), beta (15-30 Hz), and high gamma (65-200 Hz) ranges were averaged per channel. For the specparam (also known as FOOOF)^36^ method, these average curves per electrode contact for the baseline activity were processed by modelling the aperiodic curve while capturing the peaks in power (e.g. the 60 Hz noise), resulting in modelled features per power spectral curve, namely the aperiodic exponent (corresponding to the slope of the curve), the aperiodic knee (the frequency where there is a bend in the curve), and the aperiodic intercept (where the curve intercepts the y-axis). Each of these values may inform features of excitatory-inhibitory balance ^36,131,131^. Connectivity metrics involved measuring coherence and the debiased weighted phase lag index from the local field potential (LFP) as well as cross correlation using standard neural data analytical packages ^130,134,135^. To avoid bias, the neural data was processed with the analyst and code blinded to case type and recording location in that the data streams were processed in parallel without cross-comparison until the final statistical tests.

### Detecting and Mapping Interictal Discharges (IIDs)

Inter-ictal Discharges (IIDs) were automatically detected using a published algorithm that first filters the data into a 10-60 Hz band, then applies an envelope, and finally finds an appropriate threshold value modeled on a statistical distribution in the sampled envelope ^129^. We then reviewed each detection visually.

### Classification Analysis

Two support vector machine (SVM) model designs were used to test if the neural signals: 1) models used to classify the distance to the tumor or pathology and 2) models used to classify epilepsy, IDHm, and GBM diagnoses. For both model types, we subdivided the features to include largely spectral measures (aperiodic exponent, alpha, beta, and high gamma power, beta/gamma ratio, E/I slope, and IID rates per second), only connectivity measures (coherence in the theta, alpha, beta, and high gamma bands and dwPLI in the theta, alpha, beta, and high gamma bands), and using both connectivity and spectral measures.

In the first set of models to classify recordings by distance using the neural signals, an SVM pipeline with 10-fold cross-validation and probabilistic mapping was used in MATLAB. The model was trained using a Radial Basis Function (RBF) kernel with a soft-margin parameter (BoxConstraint) was constrained to 0.1, and the optimal RBF kernel variance (KernelScale) was automatically computed. The kernel scale was determined using an automated heuristic based on the median of pairwise Euclidean distances. To mitigate overfitting and prioritize a wider, more robust decision margin, the box constraint (regularization parameter) was set to 0.1. Consistent class indexing assigned Class 1 (distance <15 mm) as the positive target condition. A stratified 10-fold cross-validation scheme was deployed and Platt scaling was executed (fitPosterior) by fitting a logistic regression model onto the validation structures of each independent fold while ensuring data from individual patients are kept together in separate folds. Finally, the cross-validated discriminative capacity was evaluated with Receiver Operating Characteristic (ROC) curves via a threshold-sweeping method (perfcurve) applied to the sorted out-of-fold probability scores, mapping the cross-validated trajectory between the True Positive Rate (Sensitivity) on the Y-axis and the False Positive Rate (1 - Specificity) on the X-axis, with overall model efficacy quantified using the integrated Area Under the Curve (AUC). These calculations were done for each of the three feature sets (spectral only, connectivity only, and spectral + connectivity) per diagnosis (GBM IDHwt, IDHm, epilepsy, metastases, and all four diagnoses together or All).

In the second set of models to use features to classify diagnosis based on neural signals, we used an Error-Correcting Output Codes (ECOC) multiclass framework implemented using Support Vector Machine (SVM) binary learners. The classification pipeline proceeded through multiple steps: Data points were filtered based on quality constraints and parsed into epilepsy, GBM (IDHwt) and IDHm diagnoses to identify the predictive capacity of spectral, connectivity, and spectral + connectivity features. For each subset, features were standardized twith zscoring. Classification boundaries were mapped using a non-linear Gaussian RBF kernel. A soft-margin box constraint was fixed at 0.1 to prioritize structural generalization and minimize overfitting, and the kernel scale parameter was automatically determined via heuristic subsampling. The model was evaluated using 10-fold stratified cross validation to preserve class distribution. Generalized model performance was assessed by computing out-of-sample accuracy and aggregating out-of-fold cross-validated predictions into confusion matrices to detail specific misclassification trajectories across all evaluation classes. These calculations were done for each of the three feature sets (spectral only, connectivity only, and spectral + connectivity) per diagnosis (GBM IDHwt, IDHm, and epilepsy diagnoses).

### Statistical Analyses

Statistical comparisons were performed using non-parametric measures. There was the complication that the electrodes and tumors were in different regions and differences in anesthesia (awake vs general anesthesia). We then applied multiway ANOVA on the physiological signatures relative to brain region, tumor type, and presence of tumor. For comparisons between conditions, we further used the Kruskall-Wallis test a multiple comparisons test (Bonferoni). For groups demonstrating statistically significant differences, post-hoc pairwise comparisons were performed on the mean ranks using the MATLAB multcompare function with a Tukey-Kramer adjustment to control the family-wise error rate. These statistical analyses were conducted using MATLAB R2025b. For comparisons between two conditions, Wilcoxon rank-sum tests were performed.

## Data Availability

All data produced in the present study are available upon reasonable request to the authors

## Data and code availability

Custom code using the following packages is included in the Github repository (https://github.com/Center-For-Neurotechnology/BrainInterface3D). Open source acquisition software, RHD2000 or RHX2000 (https://intantech.com/RHX_software.html/) or OpenEphys (http://www.open-ephys.org/) ^103^ was used to record and analyze the neural data from the Intan system. Blackrock APIs (https://blackrockneurotech.com/support/) for data conversion or use of EDF reader code for import into MATLAB (https://www.mathworks.com/help/signal/ref/edfread.html) was used for the BlackRock or Natus data. Custom MATLAB code (version R2021b, R2024b. R2025a) in combination with open source code from the Fieldtrip toolbox (http://www.fieldtriptoolbox.org/), MNE python, and specparam or FOOOF (https://fooof-tools.github.io/fooof/) ^36^ was used for the majority of the analyses. Electrode tracking over the cortical surface involved 3D reconstruction of the brain surface and segmentation including vessels using 3Dslicer (https://www.slicer.org/) ^119,126^, Freesurfer 7.4 and 8.2 (https://surfer.nmr.mgh.harvard.edu/) ^108–111,118,120–124^, and the Human Connectome Project (HCP) pipeline (https://github.com/Washington-University/HCPpipelines) ^136–138^. The segmentations and surface reconstructions along with the skull was imported into Blender (https://www.blender.org/) ^104^ and electrode locations were exported using custom Python code. Geodesic distances were calculated using PyMeshLab (https://pymeshlab.readthedocs.io/en/latest/) with Python within MATLAB. The data that support the findings of this study are will be uploaded to DABI upon publication in the iEEG-BIDS format (https://bids-specification.readthedocs.io/en/stable/modality-specific-files/intracranial-electroencephalography.html and https://github.com/Center-For-Neurotechnology/BIDS_pipeline) ^128^.

De-identified neurophysiological and imaging data will be made available upon publication of this manuscript to a public repository.

## Acknowledgements

We would like to thank Yangling Chou, Melissa Murphy, Jesse Lassor, J. Scot Mackeil, and Aaron Tripp for help in data collection and, of course, the patients for their willingness to participate in this research. We would like to thank G. Rees Cosgrove and E. Antonio Chiocca for their help in gathering the data in the operating room. This research was funded to DPC, GPD, ZMW, SSC and ACP by NIH 1R01NS134410-01. RMR, AB, and MV were supported by NIH grant 1U01NS117836. DPC is also funded by R01CA307174, R01CA276765, and P50CA165962. Thin film electrode development is funded by NIH UG3NS123723, R01NS123655, and DP2-EB029757 to SAD. GPD was supported by the Paul B. and Sandra M. Edgerley MGH Research Scholar Award 2024-2029. The views and conclusions contained in this document are those of the authors and do not represent the official policies, either expressed or implied, of the funding sources.

## Contributions

Conceptualization, ACP, SSC, DPC. Methodology, ACP, SSC, JZ, ID, HR, SAD. Formal analysis, ACP, TN, JZ. Investigation, TN, BN, ID, BFC, TW, EP, ASP, RC, HKT, AB, MV, AM, HG, JCY, WM, ET, AG, JDB, DRC, AJG, PSJ, BVN, AMTR, ZMW, GPD, ST, WLB, DPC, SSC, ACP. Resources, JZ, RC, AB, MV, RMR. Data curation, ID, ACP, HR. Writing, ACP, TN, PH, ST, SSC, ASP, DPC, JZ, KLC, AJG, DRC. Visualization, ACP, TN. Supervision, ACP, SSC, DPC, GPD. Project administration, ACP, ID. Funding acquisition, ACP, SSC, SAD, ZMW, RMR, DPC, GPD.

