## Supplemental Figures and Tables for "Spatial Neurophysiological Gradients at High Resolution Differentiate Tumor Types in the Human Cortex at the Tumor-Brain Interface"

**Affiliations:**
[1] Division of Neuro-Oncology, Department of Neurosurgery, University of California San Francisco, San Francisco CA 94143, USA
[2] Department of Neurosurgery, Massachusetts General Hospital, Boston, Massachusetts 02114, USA
[3] Department of Neurology, Harvard Medical School, Massachusetts General Hospital, Boston MA 02114, USA
[4] Center for Neurotechnology and Neurorecovery, Boston MA 02114, USA
[5] Department of Neurological Surgery, Oregon Health and Science University, Portland, Oregon, 97239, USA
[6] Department of Neurosurgery, Brigham and Women's Hospital, Harvard Medical School, Massachusetts 02115, USA
[7] Department of Electrical and Computer Engineering, University of California San Diego, La Jolla, California 92093, USA
[8] Department of Neurology, Brigham and Women's Hospital, Massachusetts 02115, USA
[9] Boston VA Healthcare System, 150 South Huntington Avenue, Boston, Massachusetts 02130, USA
[10] Center for Neuro-Oncology, Department of Medical Oncology, Dana-Farber Cancer Institute, Boston MA 02215, USA

‡ Authors are co-senior authors.

*** Correspondence:**
Angelique C. Paulk


**Supplemental Table 1. Data set characteristics.**

| Broad diagnosis | Recording details | Counts |
|---|---|---|
| **Epilepsy** | Thin film circular electrode recordings | 13 |
| | Thin film linear electrode recordings | 12 |
| | 1024 thin film recordings | 2 |
| | 64 channel PMT grid | 4 |
| | 128 channel PMT grid | 3 |
| | Clinical recordings | 26 |
| | Hemisphere | Left-12, Right-22 |
| **Tumor** | Thin film circular electrode recordings | 10 |
| | Thin film linear electrode recordings | 6 |
| | 1024 thin film recordings | 5 |
| | 64 channel PMT grid | 33 |
| | 128 channel PMT grid | 12 |
| | Clinical recordings | 35 |
| | Hemisphere | Left-43, Right-19 |
| **DBS** | 64 channel PMT grid | 6 |
| | Hemisphere | Left-6, Right-0 |

**Supplemental Table 2. Diagnosis, age, and lobe and hemisphere coverage.** Participant designations include previously published designations [1–6]. Repeated designation pairs indicate separate surgical cases from the same individual: IP13 and IP34 correspond to one participant, and IP35 and IP38 correspond to another participant.

| Desig-nation | Published [illegible] | Surgical reason | Age at Surgery | Sex | Brief Diagnosis | Brain Region | Affected and Recorde |
|---|---|---|---|---|---|---|---|
| **Pt2** | | tumor | 65-70 | F | Metastasis | Parietal | Right |
| **Pt3** | IP19 | tumor | 51-55 | M | metastatic carcinoma | Temporal | Left |
| **Pt4** | Participant 15 | tumor | 65-70 | F | glioblastoma; IDH wildtype | Parietal | Left |
| **Pt5** | IP36 | epilepsy | 51-55 | F | Temporal lobe epilepsy | Temporal | Right |
| **Pt7** | | tumor | 61-65 | F | recurrent glioblastoma | Parietal | Left |
| **Pt8** | | tumor | 56-60 | F | Recurrent glioblastoma | Temporal | Right |
| **Pt9** | | tumor | 61-65 | M | astrocytoma | Frontotemporal | Right |
| **Pt10** | IP42 | tumor | 51-55 | F | Epilepsy | Occipital | Left |
| **Pt11** | IP31 | epilepsy | 51-55 | M | Post Traumatic Epilepsy | Temporal | Right |
| **Pt12** | | tumor | 51-55 | M | Astrocytoma, IDH-mutant | Frontotemporal | Right |
| **Pt13** | IP18 | epilepsy | 26-30 | M | Focal cortical Dysplasia | Frontal | Left |
| **Pt14** | | epilepsy | 55-60 | F | Epilepsy | Temporal | Left |
| **Pt15** | | tumor | 46-50 | M | Glioma | Parietal | Left |
| **Pt16** | | tumor | 56-60 | F | Metastasis | Temporal | Left |
| **Pt17** | IP17 | epilepsy | 36-40 | F | Temporal lobe epilepsy | Temporal | Left |
| **Pt18** | | epilepsy | 51-55 | M | epilepsy | Temporal | Right |
| **Pt19** | IP04 | epilepsy | 41-45 | F | Temporal lobe epilepsy | Temporal | Right |
| **Pt20** | | tumor | 71-75 | F | Glioblastoma | Frontal | Left |
| **Pt21** | IP01 | epilepsy | 31-35 | M | Temporal lobe epilepsy from previous tumor diagnosis | Temporal | Right |
| **Pt22** | | tumor | 61-65 | M | Recurrent glioblastoma | Temporal | Left |
| **Pt23** | | tumor | 76-80 | M | Metastasis | Frontal | Right |
| **Pt24** | | tumor | 41-45 | M | astrocytoma, IDH-mutant, WHO grade 3 | Frontal | Right |
| **Pt25** | IP02 | tumor | 31-35 | F | Glioma,IDH mutant | Temporal | Right |
| **Pt26** | | tumor | 51-55 | M | metastasis | Frontal | Left |
| **Pt27** | IP35, IP38 | tumor | 61-65 | F | Glioma,IDH wt | Frontal | Left |
| **Pt28** | | tumor | 66-70 | M | Glioblastoma | Frontal | Right |
| **Pt29** | | tumor | 41-45 | F | astrocytoma | Temporal | Right |
| **Pt30** | | epilepsy | 46-50 | M | Epilepsy | Temporal | Right |

| | | | | | | | |
|---|---|---|---|---|---|---|---|
| **Pt31** | IP25 | epilepsy | 31-35 | M | Hemorrhagic leukoencephalitis | Frontal | Right |
| **Pt32** | IP37 | epilepsy | 36-41 | F | Focal cortical Dysplasia | Frontal | Right |
| **Pt33** | IP15 | malformation | 51-55 | F | malformation | Temporal | Right |
| **Pt34** | IP12 | epilepsy | 36-41 | F | Temporal lobe epilepsy | Temporal | Right |
| **Pt35** | | epilepsy | 65-70 | M | Epilepsy | Frontotemporal | Right |
| **Pt36** | Participant 13 | epilepsy | 46-50 | F | lobectomy | Temporal | Right |
| **Pt37** | Participant 10 | tumor | 65-70 | M | glioma; IDH mutant | Frontal | Left |
| **Pt38** | IP13, IP34 | epilepsy | 21-25 | F | Temporal lobe epilepsy | Temporal | Right |
| **Pt39** | | tumor | 56-60 | M | metastasis | Frontal | Left |
| **Pt40** | Participant 16 | tumor | 46-50 | M | astrocytoma; IDH mutant | Frontotemporal | Left |
| **Pt42** | sub-DM1013 | movement disorder | 61-65 | M | Parkinsons Disease | Frontal | Left |
| **Pt43** | | tumor | 51-55 | M | metastasis | Parietal | Left |
| **Pt44** | IP16 | epilepsy | 41-45 | F | Temporal lobe epilepsy | Temporal | Right |
| **Pt45** | IP33 | epilepsy | 21-25 | M | Congenital insult and mesial temporal sclerosis | Temporal | Left |
| **Pt46** | IP07 | tumor | 61-65 | M | Glioma,IDH wt | Temporal | Right |
| **Pt47** | IP28 | tumor | 31-35 | F | Glioma,IDH mutant | Frontal | Right |
| **Pt48** | IP13, IP34 | epilepsy | 21-25 | F | Temporal lobe epilepsy | Temporal | Right |
| **Pt49** | | tumor | 56-60 | M | metastasis | Temporal | Left |
| **Pt50** | IP10 | epilepsy | 41-46 | M | Epilepsy due to stroke | Frontal | Left |
| **Pt51** | | tumor | 66-70 | F | glioblastoma | Frontal | Left |
| **Pt53** | sub-DM1006 | movement disorder | 61-65 | F | Dystonia | Frontal | Left |
| **Pt54** | sub-DM1005 | movement disorder | 61-65 | F | Dystonia | Frontal | Left |
| **Pt55** | Prt02 | tumor | 26-30 | F | Glioma,IDH mutant | Temporoparietal | Left |
| **Pt56** | | tumor | 46-50 | M | Recurrent glioblastoma | Frontal | Left |
| **Pt57** | | tumor | 71-75 | M | Glioma,IDH wt | Frontal | Left |
| **Pt58** | IP06 | epilepsy | 21-25 | F | Temporal lobe epilepsy from meningioma | Temporal | Left |
| **Pt60** | | tumor | 26-30 | M | glioblastoma | Frontal | Left |

| | | | | | | | |
|---|---|---|---|---|---|---|---|
| **Pt61** | IP35, IP38 | tumor | 61-65 | F | Glioma,IDH wt | Frontal | Left |
| **Pt62** | | tumor | 41-45 | M | astrocytoma | Frontal | Left |
| **Pt63** | IP03 | epilepsy | 36-40 | F | Temporal lobe epilepsy | Temporal | Right |
| **Pt64** | IP11 | tumor | 36-40 | M | Glioma,IDH mutant | Temporal | Right |
| **Pt65** | | tumor | 46-50 | F | Glioblastoma | Frontotemporal | Left |
| **Pt66** | | tumor | 31-35 | F | Astrocytoma, IDH-mutant | Insula | Right |
| **Pt67** | Participant 12 | epilepsy | 46-50 | F | cavernous malformations/ epilepsy | Temporal | Left |
| **Pt68** | IP14 | epilepsy | 26-30 | F | Focal cortical Dysplasia | Temporal | Right |
| **Pt69** | IP32 | epilepsy | 56-60 | M | Temporal lobe epilepsy | Temporal | Right |
| **Pt70** | Prt03 | tumor | 46-50 | M | Glioblastoma | Frontal | Left |
| **Pt71** | | tumor | 26-30 | F | oligodendroglioma, WHO grade 2 | Frontal | Right |
| **Pt73** | IP09 | epilepsy | 21-25 | M | Temporal lobe epilepsy | Parietal | Right |
| **Pt74** | IP23 | epilepsy | 46-70 | F | Temporal lobe epilepsy from meningioma | Temporal | Right |
| **Pt75** | | tumor | 66-70 | F | metastasis | Parietal | Left |
| **Pt76** | | tumor | 26-30 | F | IDH-mutant astrocytoma | Frontal | Left |
| **Pt77** | | tumor | 41-45 | M | Glioma | Frontal | Right |
| **Pt78** | | epilepsy | 61-65 | F | epilepsy | Frontotemporal | Right |
| **Pt79** | Participant 9 | tumor | 66-70 | M | carcinoma | Frontal | Right |
| **Pt80** | IP39 | epilepsy | 46-50 | F | Temporal lobe epilepsy | Temporal | Left |
| **Pt81** | | tumor | 61-65 | M | Glioblastoma | Parietal | Right |
| **Pt82** | HP01 | epilepsy | 26-30 | M | Neurofibromatosis Type 1 | Temporal | Left |
| **Pt83** | | tumor | 61-65 | M | recurrent glioblastoma | Temporal | Left |
| **Pt84** | IP27 | tumor | 51-55 | M | Glioma,IDH wt | Frontal | Right |
| **Pt85** | | tumor | 51-55 | F | Glioma,IDH mutant | Temporal | Left |
| **Pt86** | Participant 17 | tumor | 71-75 | F | glioblastoma; IDH wildtype | Frontotemporal | Left |
| **Pt87** | | tumor | 21-25 | F | Astrocytoma | Frontal | Right |
| **Pt88** | IP41 | epilepsy | 31-35 | F | Epilepsy | Temporal | Right |
| **Pt89** | | tumor | 76-80 | F | Glioblastoma | Temporal | Right |
| **Pt90** | | epilepsy | 26-30 | F | Temporal lobe epilepsy | Temporal | Left |
| **Pt91** | sub-DM1002 | movement disorder | 61-65 | M | Parkinsons Disease | Frontal | Left |
| **Pt92** | | tumor | 76-80 | M | glioblastoma | Temporal | Left |

| | | | | | | | |
|---|---|---|---|---|---|---|---|
| **Pt93** | | tumor | 51-55 | M | metastasis | Frontal | Left |
| **Pt94** | Prt01 | epilepsy | 56-60 | M | Temporal lobe epilepsy | Temporal | Right |
| **Pt95** | sub-DM1007 | movement disorder | 61-65 | F | Essential Tremor | Frontal | Left |
| **Pt96** | sub-DM1010 | movement disorder | 61-65 | M | Essential Tremor | Frontal | Left |
| **Pt97** | | tumor | 51-55 | F | oligodendroglioma, WHO grade 2 | Frontal | Left |
| **Pt98** | | tumor | 61-65 | M | Glioblastoma | Frontal | Left |
| **Pt99** | | tumor | 21-25 | F | Glioma | Frontal | Left |
| **Pt100** | Prt04 | tumor | 66-70 | F | Glioblastoma | Thalamus | Left |
| **Pt101** | | tumor | 66-70 | M | Glioblastoma | Temporal | Right |
| **Pt102** | | epilepsy | 56-60 | M | epilepsy | Temporal | Right |
| **Pt103** | | tumor | 76-80 | F | recurrent oligodendroglioma | Parietal | Left |
| **Pt104** | | tumor | 76-80 | M | glioblastoma | Temporal | Right |
| **Pt105** | | tumor | 46-50 | F | astrocytoma | Temporal | Left |
| **Pt106** | | tumor | 56-60 | F | glioblastoma | Frontal | Left |
| **Pt107** | | tumor | 41-45 | F | recurrent oligodendroglioma | Parietal | Right |

**Supplemental Table 3. Distribution of recording devices and concurrent clinical electrodes across cases.** Recording devices included PEDOT:PSS microelectrodes with multiple device designs. X channel strip electrodes denote X-contact strip electrodes with 1 cm spacing.

| Desig-nation | Surgical reason | Surgical reason | Research electrodes | Electrode layout | Clinical electrodes | Photo or video |
|---|---|---|---|---|---|---|
| **Pt2** | tumor | not awake | PMT grid | Electrode_design | Clinical_electrode_recording | Yes |
| **Pt3** | tumor | awake | PEDOT:PSS | Circular grid | 4 channel strip | Yes |
| **Pt4** | tumor | awake | PNtRs | 32 x 32, 1024 ch | 64 channel grid | Yes |
| **Pt5** | epilepsy | awake | PEDOT:PSS | 50 micron pitch 2 column | 8 channel strip | Yes |
| **Pt7** | tumor | not awake | none | none | 4 channel strip | Yes |
| **Pt8** | tumor | not awake | PMT grid | 64 channel, long | 4 channel strip, 4 channel strip | Yes |
| **Pt9** | tumor | not awake | PMT grid | 64 channel, long | 8 channel strip | Yes |
| **Pt10** | tumor | not awake | PEDOT:PSS | 50 micron pitch 2 column | none | Yes |
| **Pt11** | epilepsy | not awake | PEDOT:PSS | Circular grid | 8 channel strip | Yes |
| **Pt12** | tumor | not awake | PMT grid | 64 channel, long | none | Yes |
| **Pt13** | epilepsy | not awake | PEDOT:PSS | Circular grid | 8 channel strip | Yes |
| **Pt14** | epilepsy | not awake | PMT grid | 64 channel, long | none | Yes |
| **Pt15** | tumor | not awake | none | none | 8 channel strip | Yes |
| **Pt16** | tumor | awake | none; 6 channel strip | none; 6 channel strip | 6 channel strip | Yes |
| **Pt17** | epilepsy | not awake | PEDOT:PSS | Circular grid | 4 channel strip, 6 channel depth | Yes |
| **Pt18** | epilepsy | not awake | PMT grid | 64 channel, long | none | Yes |
| **Pt19** | epilepsy | not awake | PEDOT:PSS | 50 micron pitch 2 column | 4 channel strip, 8 channel depth, 4 channel depth | Yes |
| **Pt20** | tumor | not awake | PMT grid | 64 channel, long | none | Yes |
| **Pt21** | epilepsy | not awake | PEDOT:PSS | 50 micron pitch 2 column | 6 channel strip, 6 contact depth | No |
| **Pt22** | tumor | awake | PMT grid | 64 channel, long | none | Yes |

| | | | | | | |
|---|---|---|---|---|---|---|
| **Pt23** | tumor | not awake | PMT grid | 64 channel, long | none | Yes |
| **Pt24** | tumor | not awake | PMT grid | 128 channel, rectangular | 6 channel strip | Yes |
| **Pt25** | tumor | awake | PEDOT:PSS | 50 micron pitch 2 column | 6 channel strip | No |
| **Pt26** | tumor | awake | none | none | 8 channel strip, 4 channel strip | Yes |
| **Pt27** | tumor | awake | PEDOT:PSS | 50 micron pitch 2 column | 8 channel strip, depth | Yes |
| **Pt28** | tumor | not awake | PMT grid | 64 channel, long | 4 channel strip | Yes |
| **Pt29** | tumor | not awake | PMT grid | 64 channel, long | none | Yes |
| **Pt30** | epilepsy | not awake | PMT grid | 128 channel, rectangular | 4 channel strip, 4 channel strip | Yes |
| **Pt31** | epilepsy | not awake | PEDOT:PSS | Circular grid | 8 channel strip | Yes |
| **Pt32** | epilepsy | awake | PEDOT:PSS | Circular grid | 8 channel strip, 4 channel strip | Yes |
| **Pt33** | malformation | awake | PEDOT:PSS | 50 micron pitch 2 column | 8 channel strip | Yes |
| **Pt34** | epilepsy | awake | PEDOT:PSS | 50 micron pitch 2 column | 6 channel strip | Yes |
| **Pt35** | epilepsy | not awake | none | none | 64 channel Adtech | Yes |
| **Pt36** | epilepsy | not awake | PNtRs | 32 x 32, 1024 ch | 64 channel grid | Yes |
| **Pt37** | tumor | awake | PNtRs | 32 x 32, 1024 ch | 64 channel grid | Yes |
| **Pt38** | epilepsy | not awake | PEDOT:PSS | 50 micron pitch 2 column | none | Yes |
| **Pt39** | tumor | not awake | Layer 7; PMT grid | Layer7; 64 channel, long | 6 channel strip | Yes |
| **Pt40** | tumor | awake | PNtRs | 32 x 32, 1024 ch | 64 channel grid | Yes |
| **Pt42** | movement disorder | awake | PMT grid | 64 channel, long | DBS electrode | Yes |
| **Pt43** | tumor | not awake | PMT grid | 64 channel, long | 8 channel strip | Yes |
| **Pt44** | epilepsy | not awake | PEDOT:PSS | 50 micron pitch 2 column | 4 channel strip, 4 channel depth | Yes |
| **Pt45** | epilepsy | not awake | PEDOT:PSS | Circular grid | 64 channel grid | Yes |
| **Pt46** | tumor | awake | PEDOT:PSS | 50 micron pitch 2 column | 8 channel strip | Yes |
| **Pt47** | tumor | awake | PEDOT:PSS | Circular grid | 8 channel strip | Yes |
| **Pt48** | epilepsy | not awake | PEDOT:PSS | Circular grid | 16 channel grid (4 x 4) | Yes |

| | | | | | | |
|---|---|---|---|---|---|---|
| **Pt49** | tumor | awake | none | none | 8 channel strip | Yes |
| **Pt50** | epilepsy | not awake | PEDOT:PSS | 50 micron pitch 2 column | 6 channel strip, 6 channel strip | Yes |
| **Pt51** | tumor | awake | PMT grid; none | 128 channel, rectangular | 6 channel strip | Yes |
| **Pt53** | movement disorder | awake | PMT grid | 64 channel, long | DBS electrode | Yes |
| **Pt54** | movement disorder | awake | PMT grid | 64 channel, long | DBS electrode | Yes |
| **Pt55** | tumor | awake | none | none | 8 channel strip | No |
| **Pt56** | tumor | awake | none | none | 64 channel Adtech | Yes |
| **Pt57** | tumor | not awake | none | none | 6 channel strip | Yes |
| **Pt58** | epilepsy | not awake | PEDOT:PSS | 50 micron pitch 2 column | 8 channel strip, 20 channel grid | Yes |
| **Pt60** | tumor | awake | none | none | 8 channel strip | Yes |
| **Pt61** | tumor | awake | PEDOT:PSS | 50 micron pitch 2 column | none | Yes |
| **Pt62** | tumor | awake | none | none | 8 channel strip | Yes |
| **Pt63** | epilepsy | not awake | PEDOT:PSS | 50 micron pitch 2 column | 8 channel strip, 8 channel depth, 8 channel depth | Yes |
| **Pt64** | tumor | not awake | PEDOT:PSS | 50 micron pitch 2 column | 8 channel strip | Yes |
| **Pt65** | tumor | awake | PMT grid | 128 channel, rectangular | none | Yes |
| **Pt66** | tumor | not awake | PMT grid | 64 channel, long | none | Yes |
| **Pt67** | epilepsy | not awake | PNtRs | 32 x 32, 1024 ch | 64 channel grid | Yes |
| **Pt68** | epilepsy | not awake | PEDOT:PSS | 50 micron pitch 2 column | 4 channel strip, 4 channel depth | Yes |
| **Pt69** | epilepsy | not awake | PEDOT:PSS | Circular grid | 8 channel strip | Yes |
| **Pt70** | tumor | awake | nonenone | nonenone | 8 channel strip | No |
| **Pt71** | tumor | not awake | PMT grid | 64 channel, long | none | Yes |
| **Pt73** | epilepsy | not awake | PEDOT:PSS | 50 micron pitch 2 column | 6 channel strip, 6 channel strip | Yes |
| **Pt74** | epilepsy | not awake | PEDOT:PSS | Circular grid | none | Yes |
| **Pt75** | tumor | not awake | PMT grid | 64 channel, long | 8 channel strip | Yes |
| **Pt76** | tumor | not awake | PMT grid | 64 channel, long | none | Yes |

| | | | | | | |
|---|---|---|---|---|---|---|
| **Pt77** | tumor | not awake | PMT grid | 64 channel, long | none | Yes |
| **Pt78** | epilepsy | not awake | PMT grid | 64 channel, long | none | Yes |
| **Pt79** | tumor | awake | PNtRs | 32 x 32, 1024 ch | none | Yes |
| **Pt80** | epilepsy | awake | PEDOT:PSS | PEDOT short depth | 8 channel strip, 4 channel strip | Yes |
| **Pt81** | tumor | not awake | PMT grid | 128 channel, rectangular | 8 channel strip | Yes |
| **Pt82** | epilepsy | not awake | PEDOT:PSS | PEDOT short depth | 6 channel strip | Yes |
| **Pt83** | tumor | awake | PMT grid | 64 channel, long | 8 channel strip | Yes |
| **Pt84** | tumor | awake | PEDOT:PSS | 50 micron pitch 2 column | 8 channel strip | Yes |
| **Pt85** | tumor | awake | nonenone | nonenone | 6 channel strip | Yes |
| **Pt86** | tumor | awake | PNtRs | 16 x 64, 1024 ch | 64 channel grid | Yes |
| **Pt87** | tumor | not awake | PMT grid | 64 channel, long | 8 channel strip | Yes |
| **Pt88** | epilepsy | not awake | PEDOT:PSS | 50 micron pitch 2 column | 4 channel strip, 4 channel depth, 6 channel depth | Yes |
| **Pt89** | tumor | not awake | 6 channel strip | 6 channel strip | none | Yes |
| **Pt90** | epilepsy | not awake | PMT grid | 64 channel, long | none | Yes |
| **Pt91** | movement disorder | awake | PMT grid | 64 channel, long | DBS electrode | Yes |
| **Pt92** | tumor | not awake | PMT grid | 64 channel, long | none | Yes |
| **Pt93** | tumor | not awake | PMT grid | 64 channel, long | 8 channel strip | Yes |
| **Pt94** | epilepsy | not awake | none | none | 8 channel strip, 4 channel depth, 8 channel depth | No |
| **Pt95** | movement disorder | awake | PMT grid | 64 channel, long | DBS electrode | Yes |
| **Pt96** | movement disorder | awake | PMT grid | 64 channel, long | DBS electrode | Yes |
| **Pt97** | tumor | awake | PMT grid | 64 channel, long | 8 channel strip | Yes |
| **Pt98** | tumor | not awake | PMT grid | 64 channel, long | none | Yes |
| **Pt99** | tumor | not awake | PMT grid | 64 channel, long | none | Yes |
| **Pt100** | tumor | awake | none | none | 8 channel strip | No |

| | | | | | | |
|---|---|---|---|---|---|---|
| **Pt101** | tumor | not awake | PMT grid | 64 channel, long | none | Yes |
| **Pt102** | epilepsy | not awake | PMT grid | 128 channel, rectangular | 4 channel strip, 4 channel strip | Yes |
| **Pt103** | tumor | not awake | PMT grid | 64 channel, long | none | Yes |
| **Pt104** | tumor | not awake | PMT grid | 64 channel, long | none | Yes |
| **Pt105** | tumor | not awake | PMT grid | 64 channel, long | none | Yes |
| **Pt106** | tumor | not awake | PMT grid | 64 channel, long | none | Yes |
| **Pt107** | tumor | awake | PMT grid | 64 channel, long | none | Yes |

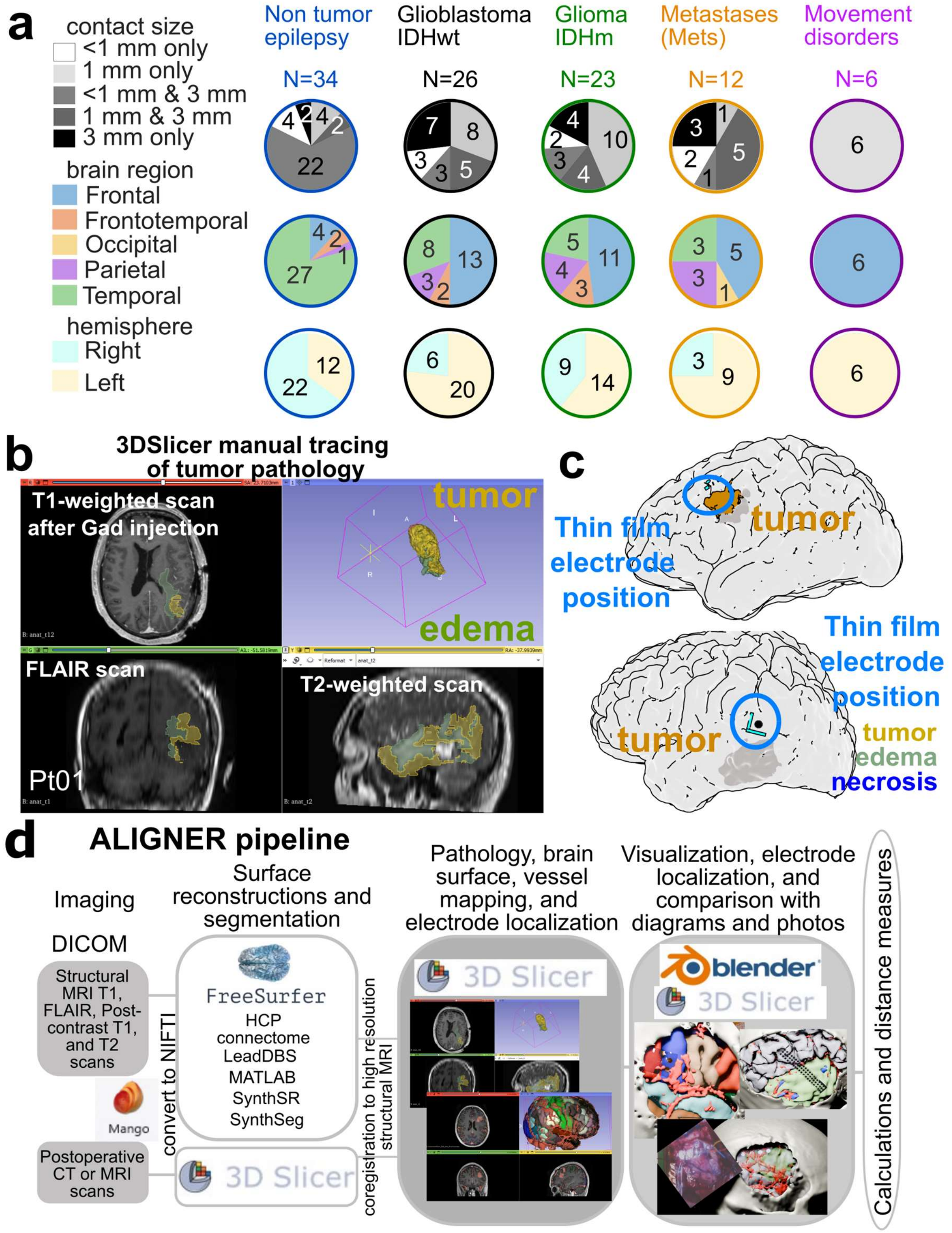

**Supplemental Figure 1. Demographics and Pipeline for localizing electrodes on the brain.** **a.** Electrode and brain region sampling information across participants. Distribution of types and numbers of cases, brain regions covered, and hemisphere of recording involving clinical ECoG and meso-ECoG and µECoG between the diagnoses, color coded as above and as in the legend. **b-d.** Advanced Localization and Imaging Guidance for Neurosurgical Electrode Recording, ALIGNER.

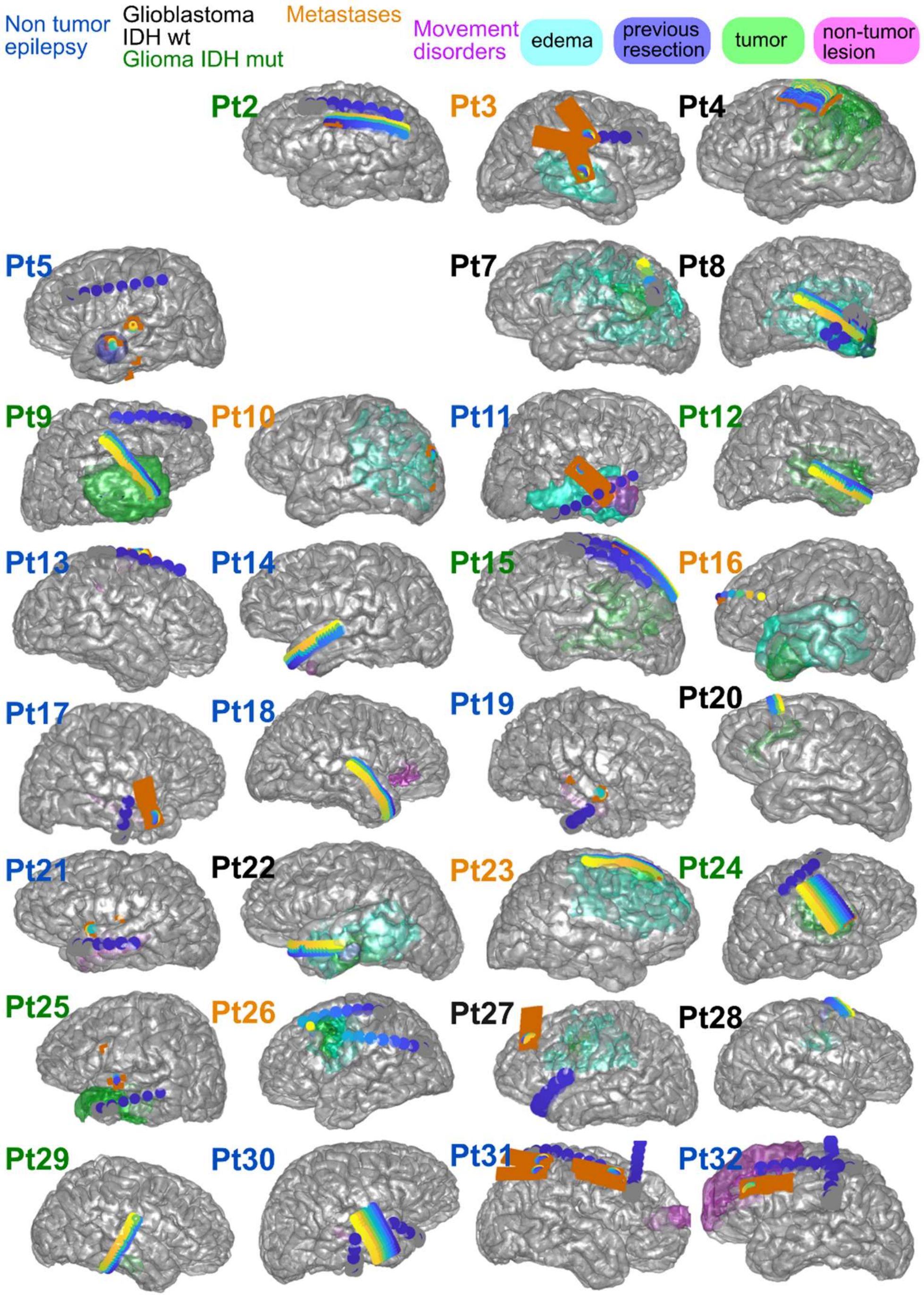

**Supplemental Figure 2. Locations of electrodes for individual participants relative to identified pathologies.** Electrodes are indicated by blue to yellow dots (color coded to indicate channel number). PtXXX designations are per participant, with the color of the text indicating the diagnosis: non-tumor epilepsy cases (blue), IDH wt glioblastomas (black), IDH mutant gliomas (green), metastases (orange), other tumor diagnoses (cyan), and movement disorders (magenta). Grey surfaces indicate the surface of the brain and color coded surfaces indicate non-tumor lesion (magenta), previous resection (dark blue), edema (cyan), and tumor (green).

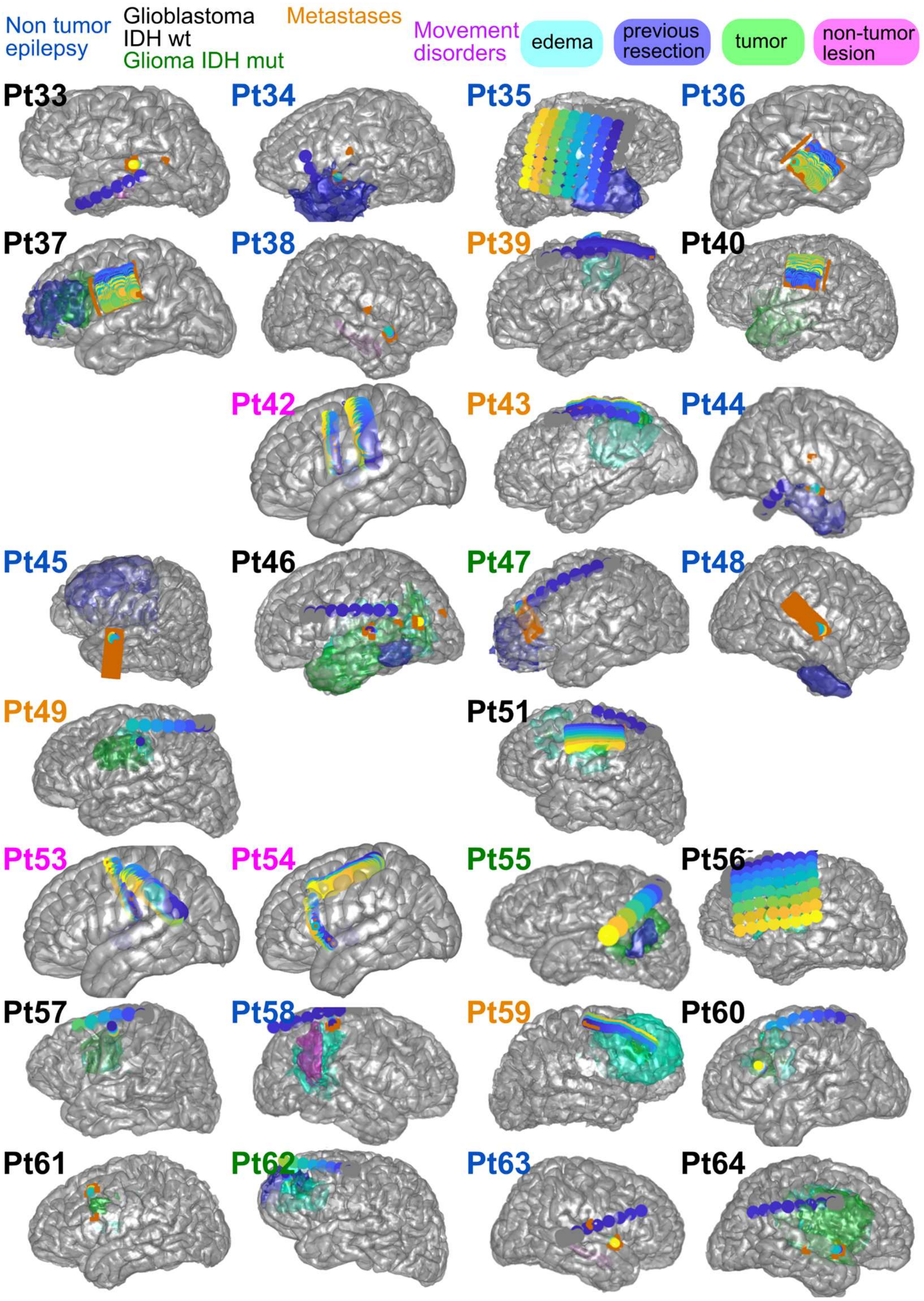

**Supplemental Figure 3. Locations of electrodes for individual participants relative to identified pathologies.** Electrodes are indicated by blue to yellow dots (color coded to indicate channel number). PtXXX designations are per participant, with the color of the text indicating the diagnosis: non-tumor epilepsy cases (blue), IDH wt glioblastomas (black), IDH mutant gliomas (green), metastases (orange), other tumor diagnoses (cyan), and movement disorders (magenta). Grey surfaces indicate the surface of the brain and color coded surfaces indicate non-tumor lesion (magenta), previous resection (dark blue), edema (cyan), and tumor (green).

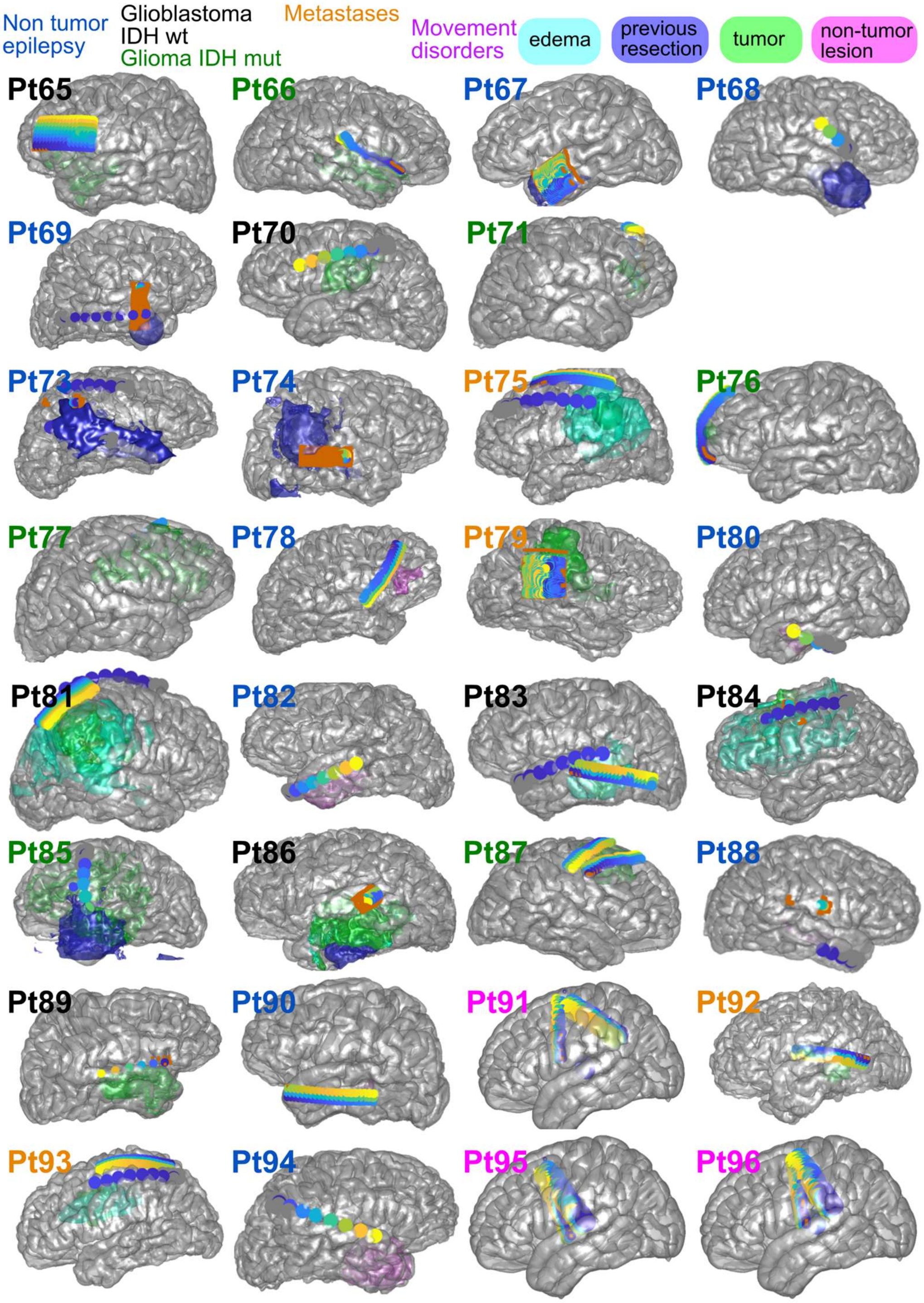

**Supplemental Figure 4. Locations of electrodes for individual participants relative to identified pathologies.** Electrodes are indicated by blue to yellow dots (color coded to indicate channel number). PtXXX designations are per participant, with the color of the text indicating the diagnosis: non-tumor epilepsy cases (blue), IDH wt glioblastomas (black), IDH mutant gliomas (green), metastases (orange), other tumor diagnoses (cyan), and movement disorders (magenta). Grey surfaces indicate the surface of the brain and color-coded surfaces indicate non-tumor lesion (magenta), previous resection (dark blue), edema (cyan), and tumor (green).

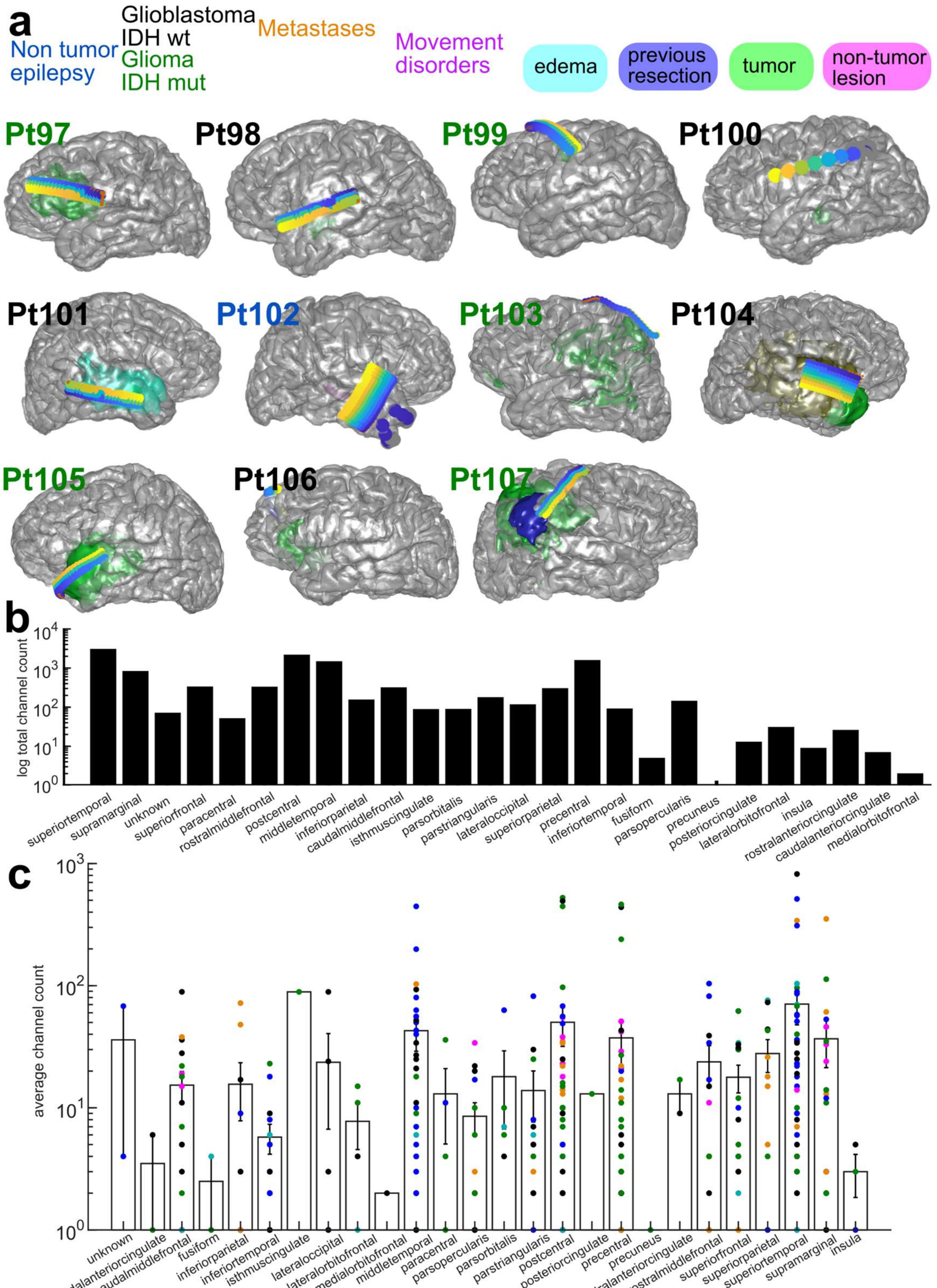

**Supplemental Figure 5. Locations of electrodes for individual participants relative to identified pathologies and across brain regions. a**. Electrodes are indicated by blue to yellow dots (color coded to indicate channel number). PtXXX designations are per participant, with the color of the text indicating the diagnosis: non-tumor epilepsy cases (blue), IDH wt glioblastomas (black), IDH mutant gliomas (green), metastases (orange), other tumor diagnoses (cyan), and movement disorders (magenta). Grey surfaces indicate the surface of the brain and color-coded surfaces indicate non-tumor lesion (magenta), previous resection (dark blue), edema (cyan), and tumor (green). **b.** Total counts of electrodes per brain region (DKT atlas), N=107. **c.** Average counts of electrodes per brain regions, color coded to diagnosis, N=107.

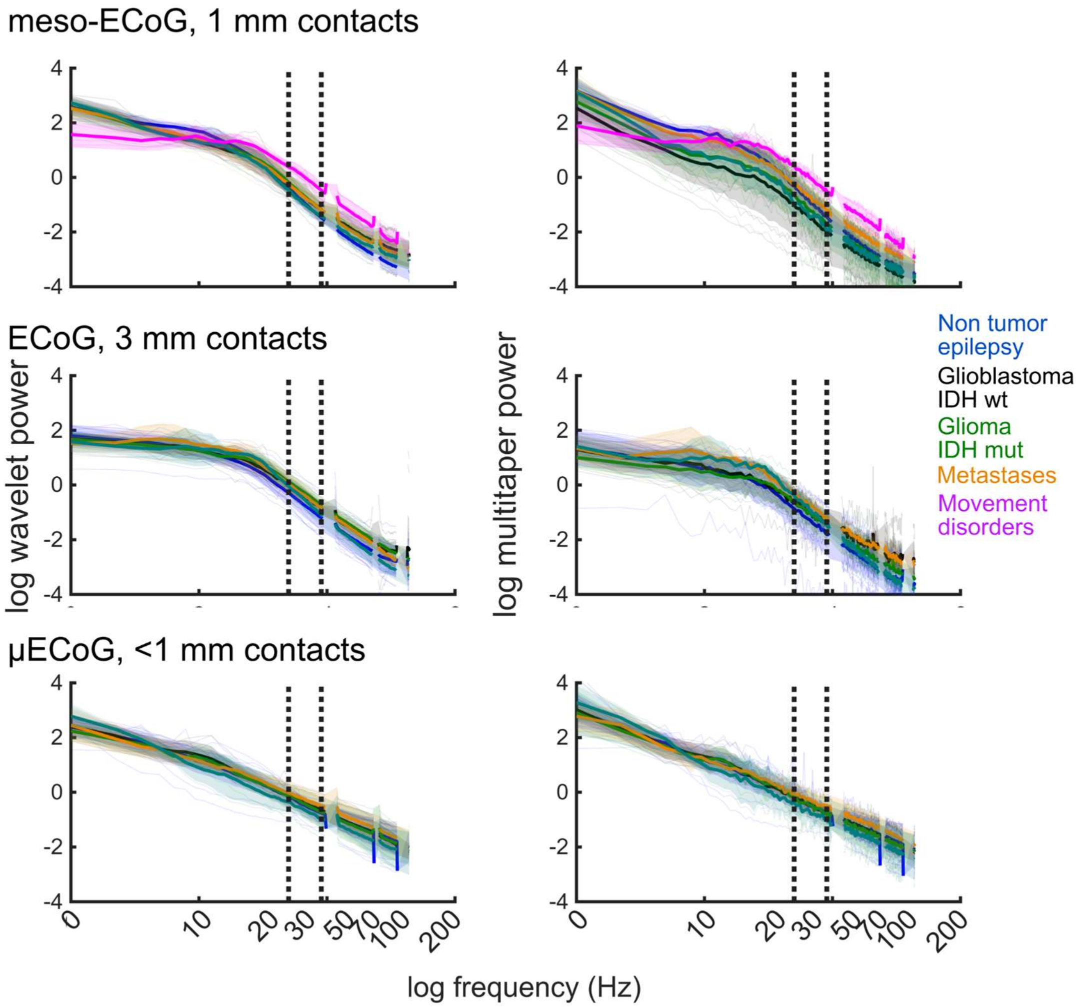

**Supplemental Figure 6. Power spectral curves across the data set and diagnoses, N=101.** Power spectral curves calculated using wavelet analyses (left) and multitaper analyses (right) averaged per device across channels per participant (thin lines) and averaged across diagnoses (thick line) with shaded error bars indicating the standard error across the data set per frequency step, with top from the meso-ECoG electrodes, middle from the clinical ECoG electrodes, and the bottom curves from the μECoG contacts.

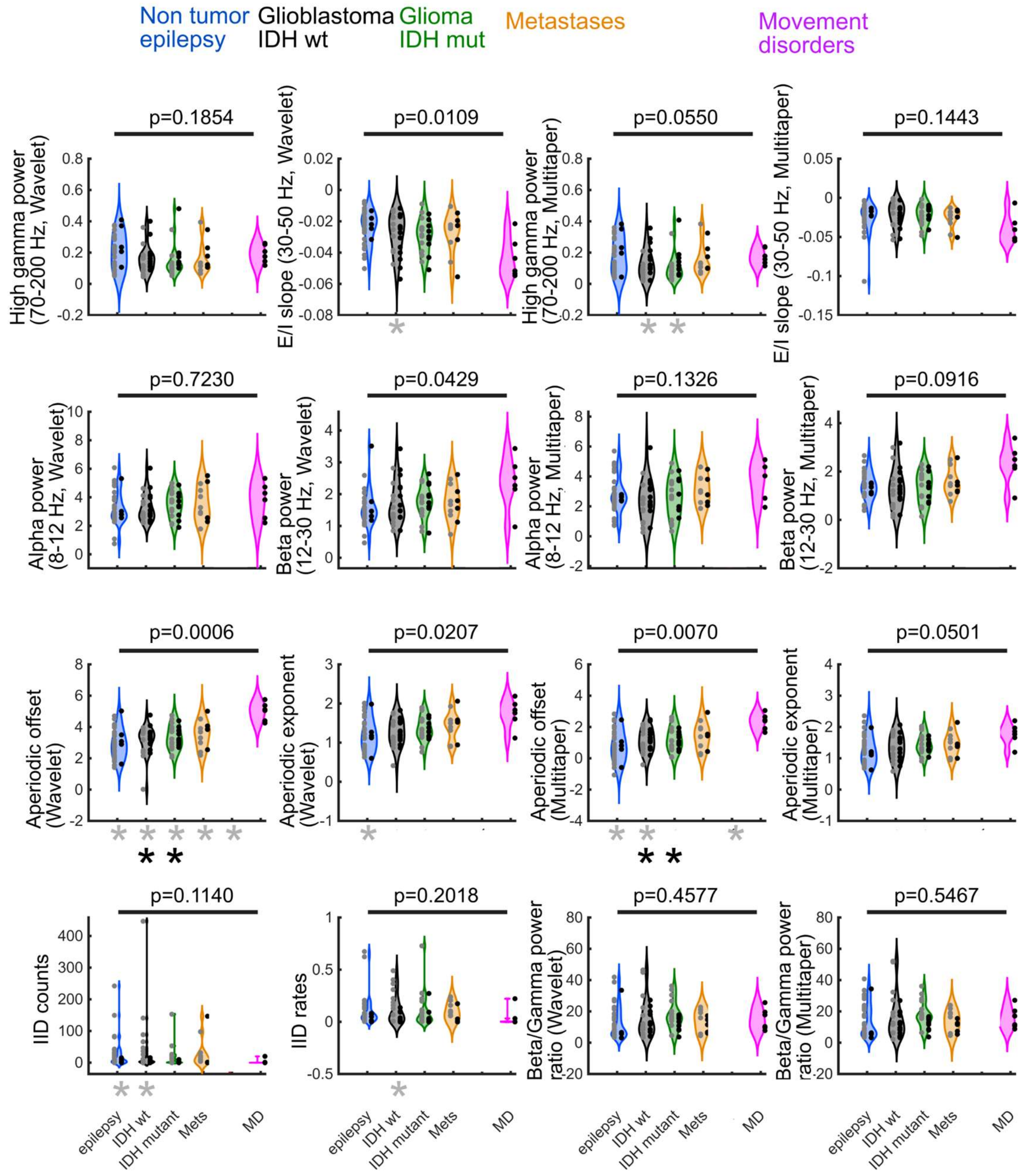

**Supplemental Figure 7. Average values per diagnosis for multiple metrics on the per-participant level, N=101**. Violin plots of per-participant-level average evaluated metrics included local field potential (LFP) recording power analysis at high gamma power, alpha power, and beta power, the excitatory/inhibitory slope of the spectral curve between 30 and 50 Hz (E/I slope), the aperiodic exponent, knee, and offset (FOOOF measures) for the wavelet and multitaper power spectral estimates. Black dots are from cases when the participants were awake under monitored anesthesia care (MAC) and grey dots are from when the participants were under general anesthesia. Black asterisks- significantly different from movement disorder recordings for all recordings under monitored anesthesia care (MAC, awake). Grey asterisks- significantly different from movement disorder (MD) recordings for all recordings under general anesthesia (GA, not awake). GBM-glioblastoma; IDHm- isocitrate dehydrogenase (IDH) mutation; IDHwt-IDH wild type; Mets-metastases; MD- movement disorder.

**E**. Violin plots of per-participant-level average clinical ECoG activity high gamma power (HGP) and aperiodic exponent.. Black dots are from cases when the participants were awake under monitored anesthesia care (MAC) and grey dots are from when the participants were under general anesthesia. Grey asterisk- significantly different from movement disorder (MD) recordings for all recordings under general anesthesia (GA, not awake).

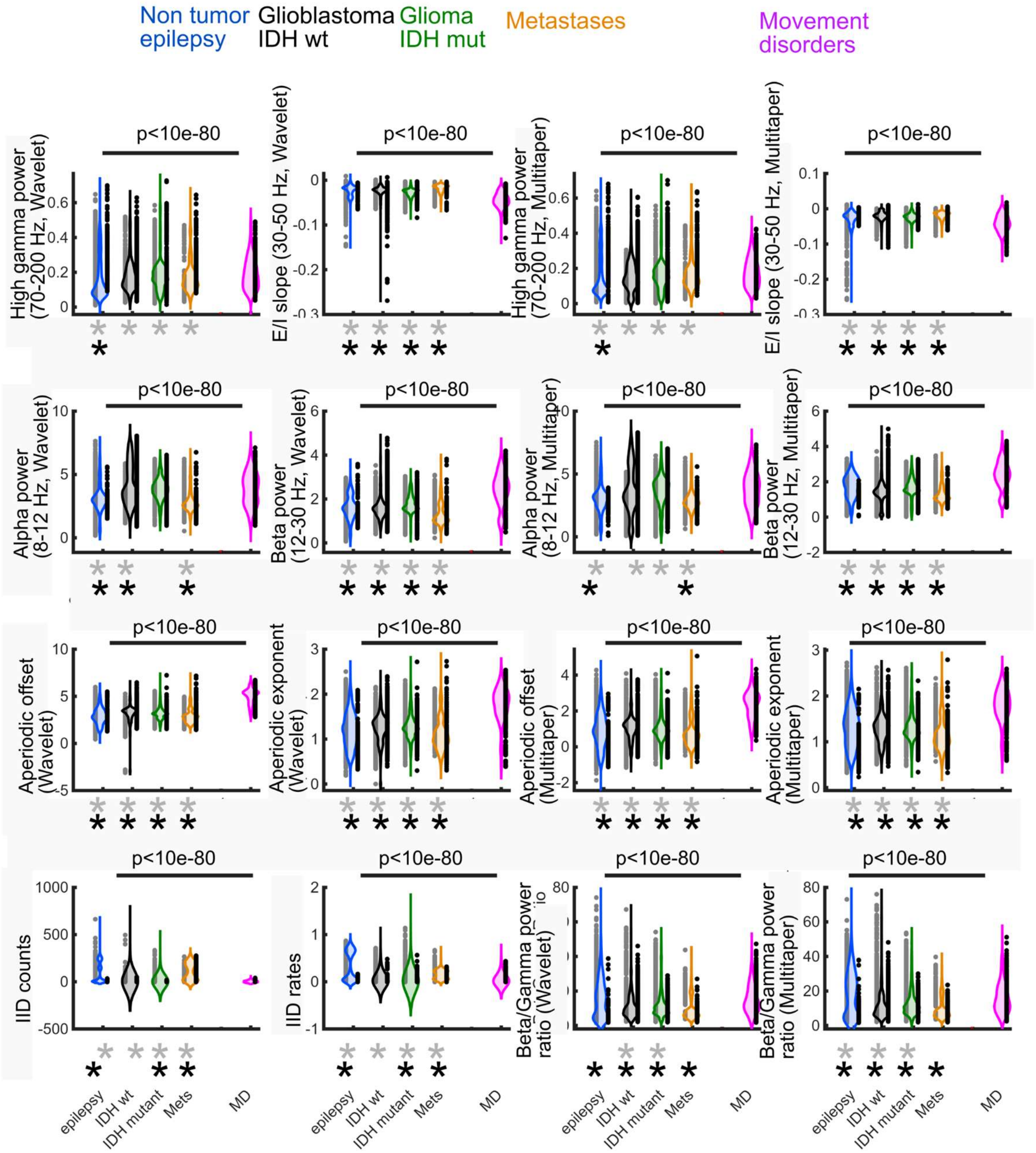

**Supplemental Figure 8. Average values per diagnosis for multiple metrics on the per-contact level, N=101.** Violin plots of per-contact-level average evaluated metrics included local field potential (LFP) recording power analysis at high gamma power, alpha power, and beta power, the excitatory/inhibitory slope of the spectral curve between 30 and 50 Hz (E/I slope), the aperiodic exponent (FOOOF measure) for the wavelet and multitaper power spectral estimates. Black dots are from cases when the participants were awake under monitored anesthesia care

(MAC) and grey dots are from when the participants were under general anesthesia. Black asterisks- significantly different from movement disorder recordings for all recordings under monitored anesthesia care (MAC, awake). Grey asterisks- significantly different from movement disorder (MD) recordings for all recordings under general anesthesia (GA, not awake). GBM- glioblastoma; IDHm- isocitrate dehydrogenase (IDH) mutation; IDHwt-IDH wild type glioblastoma; Mets-metastases; MD- movement disorder.

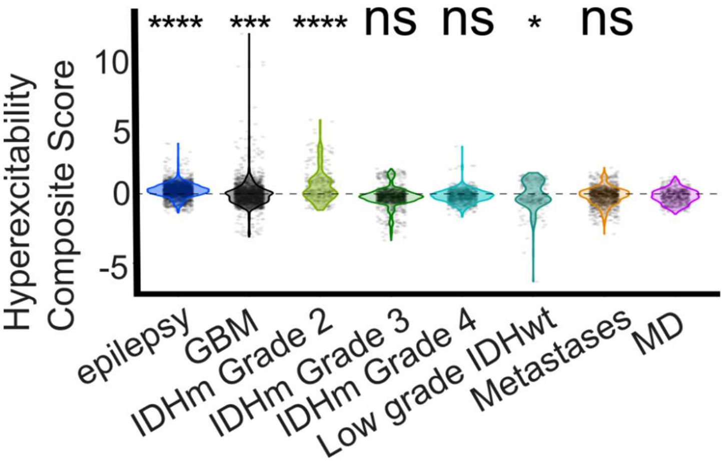

**Supplemental Figure 9. Hyperexcitability PCA analysis.** Composite hyperexcitability score. p-values are from Kruskal-Wallis multiple comparisons tests. Statistical comparisons are between MD and other diagnoses. GBM-glioblastoma; IDHm- isocitrate dehydrogenase (IDH) mutation; IDHwt-IDH wild type; Mets-metastases; MD- movement disorder.

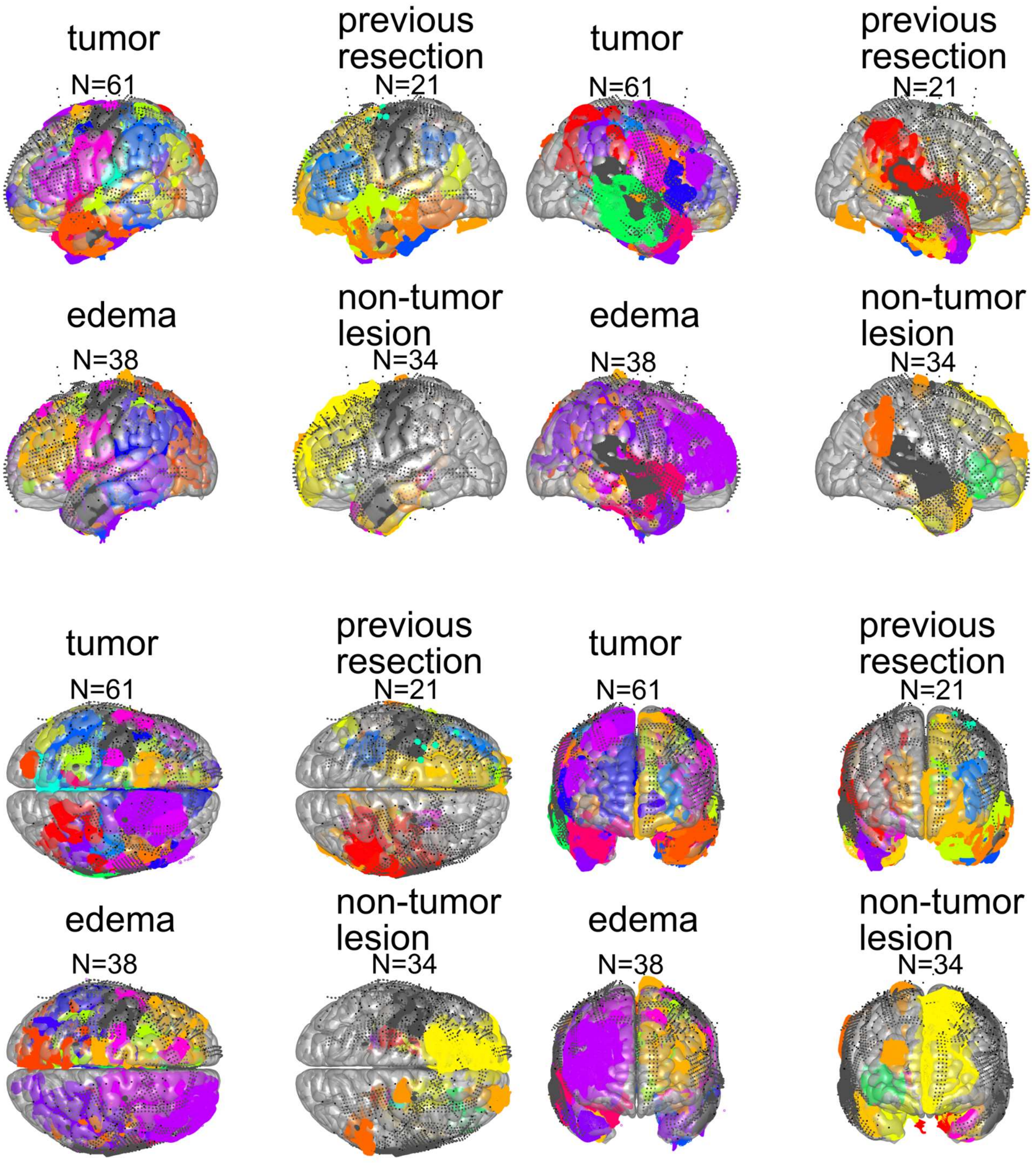

**Supplemental Figure 10. Pathologies mapped to MNI brain locations across the data set.** Different colors of the pathology-related scatter plot indicate different participants.

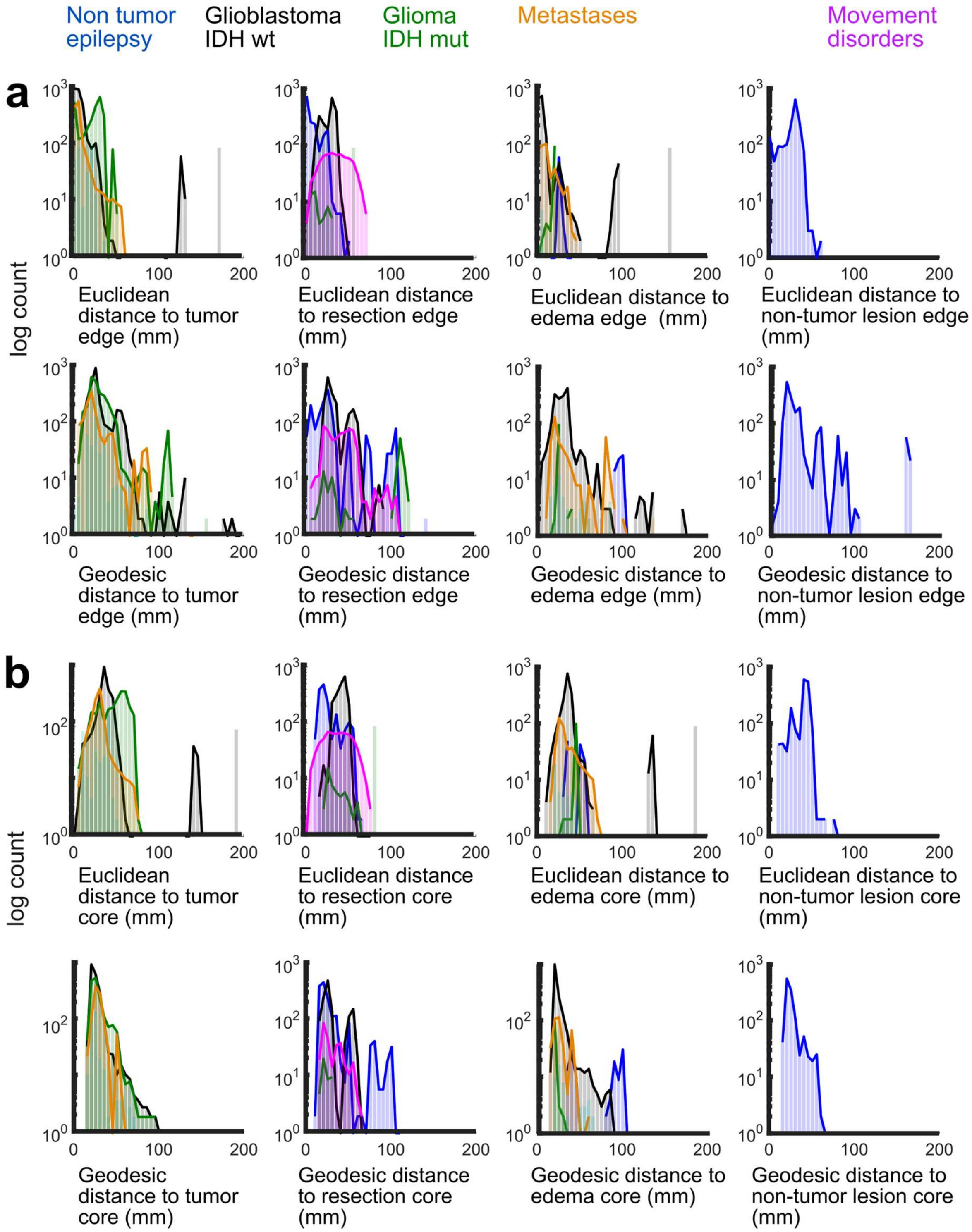

**Supplemental Figure 11. Distribution of distances between electrode contacts and, where appropriate, tumor, edema, previous resection, and non-tumor lesions (N=101). a-b.** Euclidean (**a**) and Geodesic (**b**) distance distributions. The previous resection for the movement disorder group was the location of the inserted DBS lead into the burr hole (to act as a control).

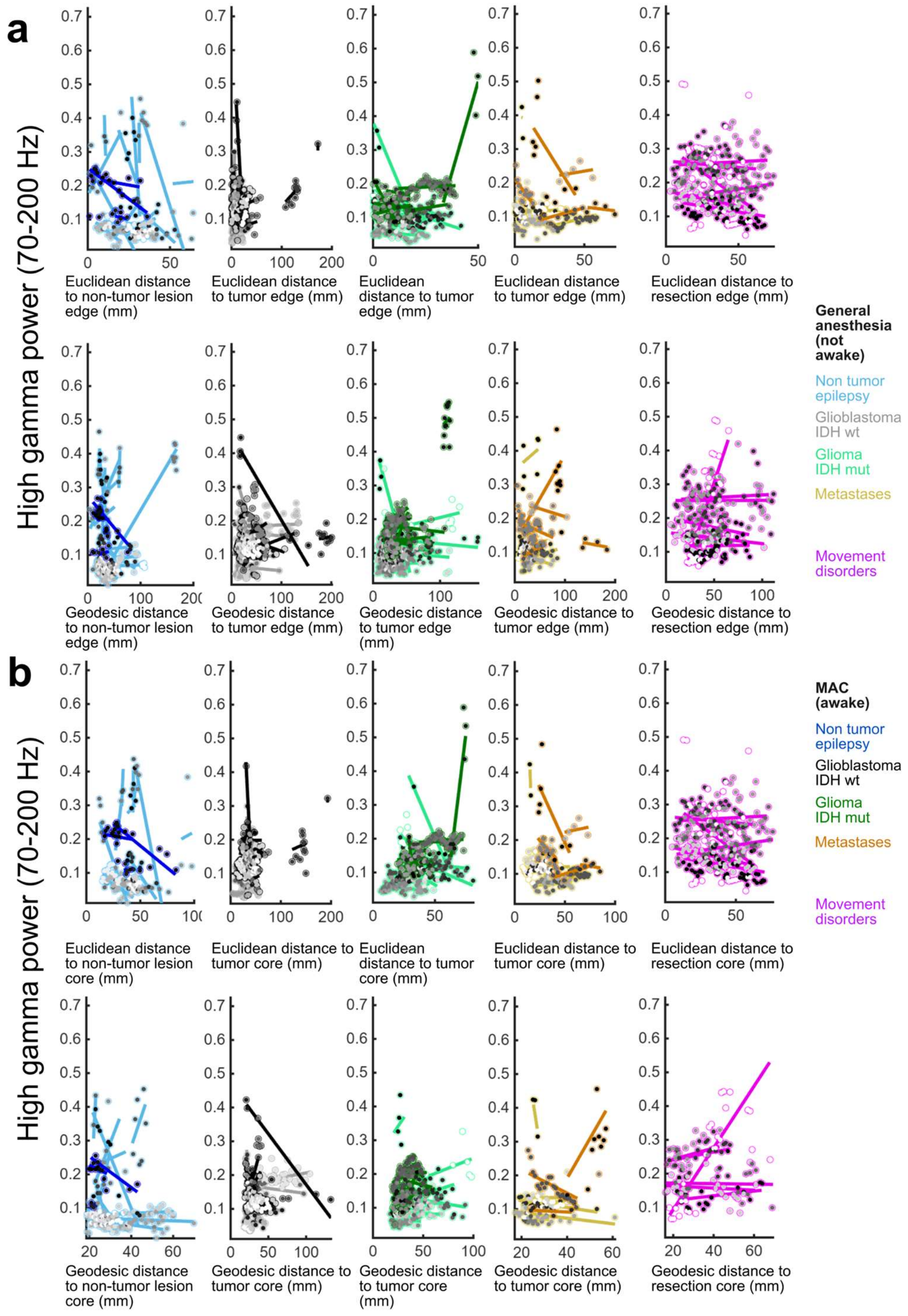

**Supplemental Figure 12. Distance to the pathology versus high gamma power across diagnoses**. **A**. Euclidean and Geodesic distances to the pathology edge to contacts per device per participant (distances and neural measures averaged) for high gamma power. **B**. Euclidean and Geodesic distances to the pathology core to contacts per device per participant (distances and neural measures averaged) for high gamma power. Lines indicate significant correlations per participant.

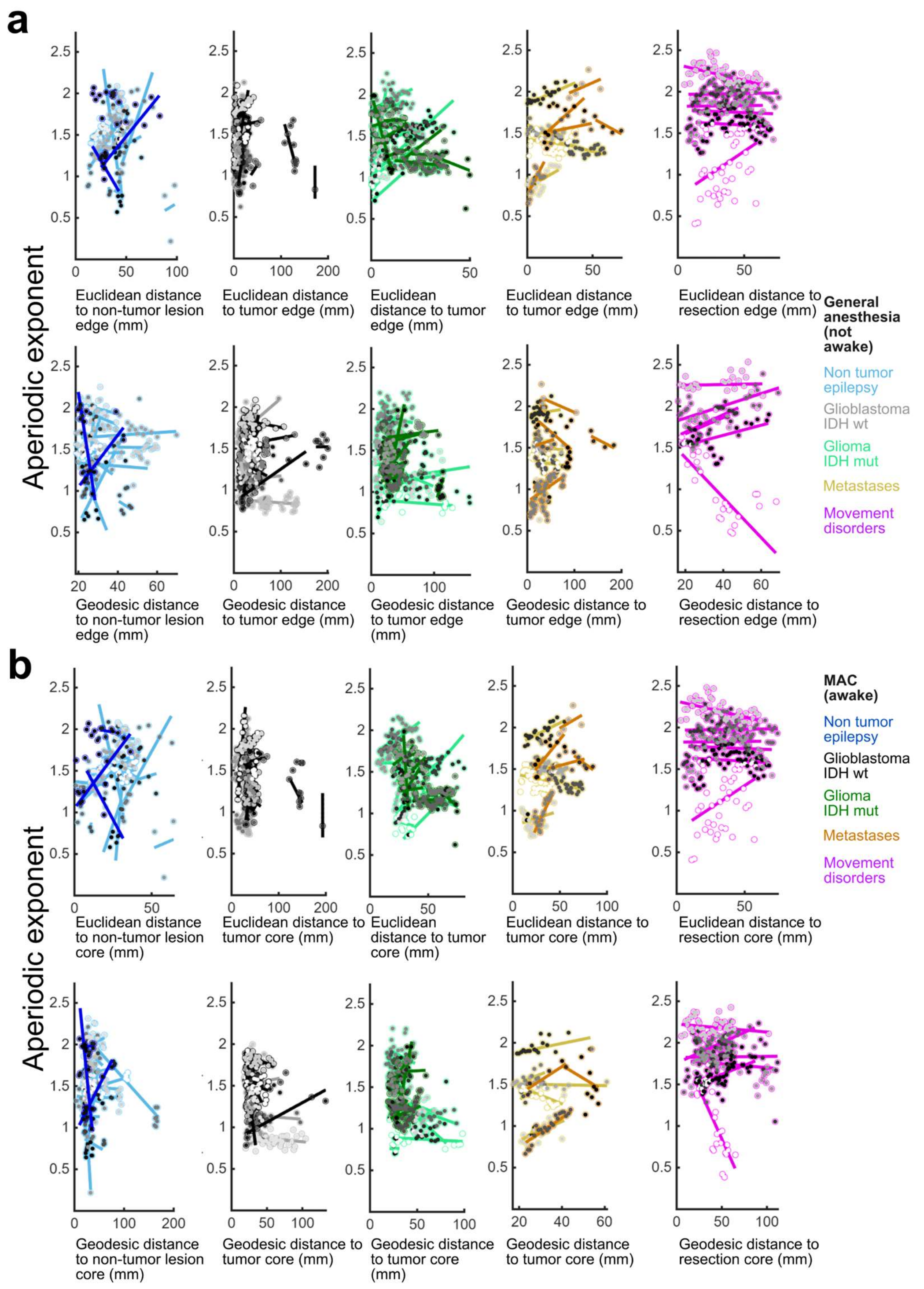

**Supplemental Figure 13. Distance to the pathology versus aperiodic exponent across diagnoses**. **A**. Euclidean and Geodesic distances to the pathology edge to contacts per device per participant (distances and neural measures averaged) for aperiodic exponent (wavelet). **B**. Euclidean and Geodesic distances to the pathology core to contacts per device per participant (distances and neural measures averaged) for aperiodic exponent (wavelet). Lines indicate significant correlations per participant.

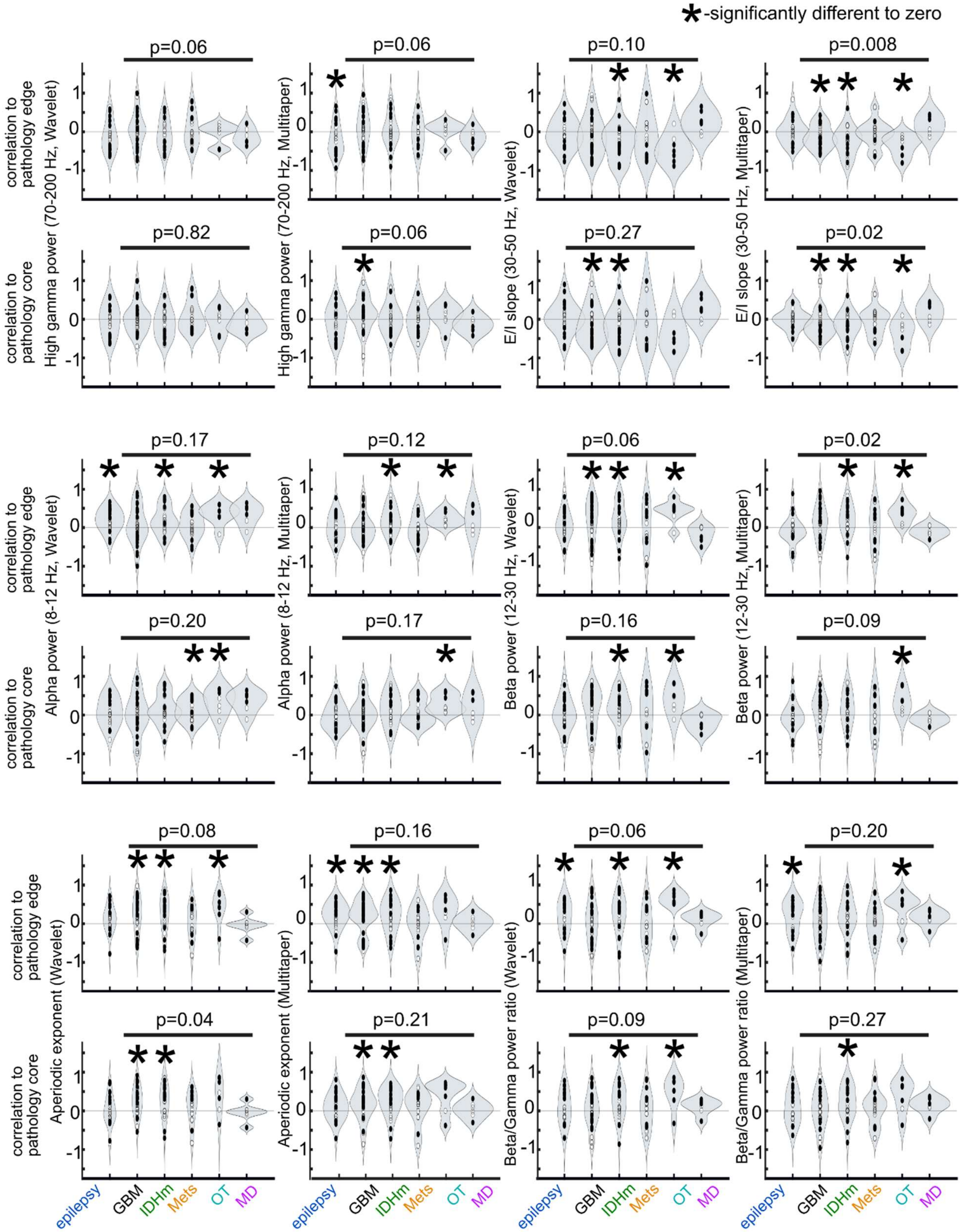

**Supplemental Figure 14. Correlation values between Euclidean distance to the pathology (tumor, lesion, edema, or resection) edge or core and neurophysiological measures.**

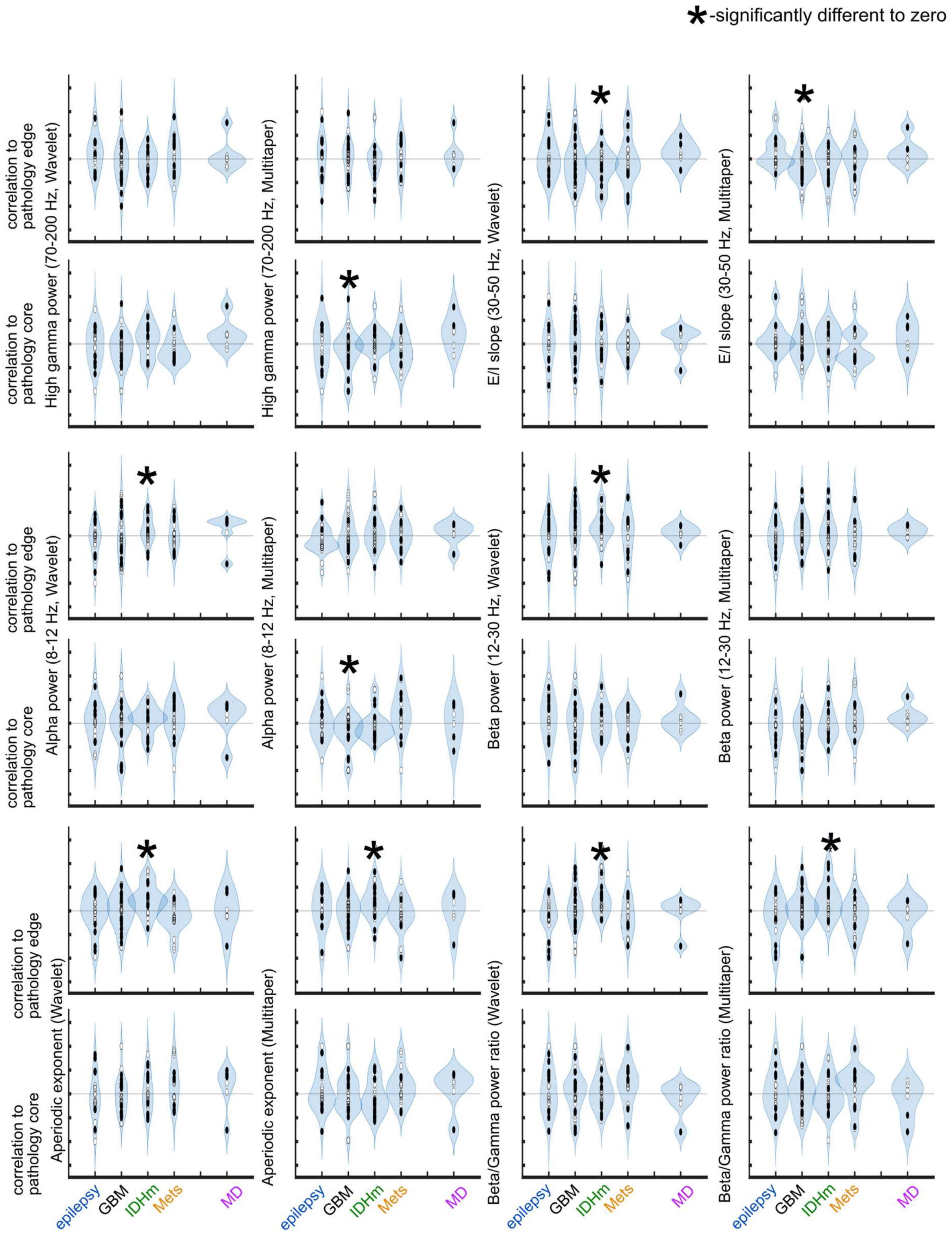

**Supplemental Figure 15. Correlation values between Geodesic distance to the pathology (tumor, lesion, edema, or resection) edge or core and neurophysiological measures.**

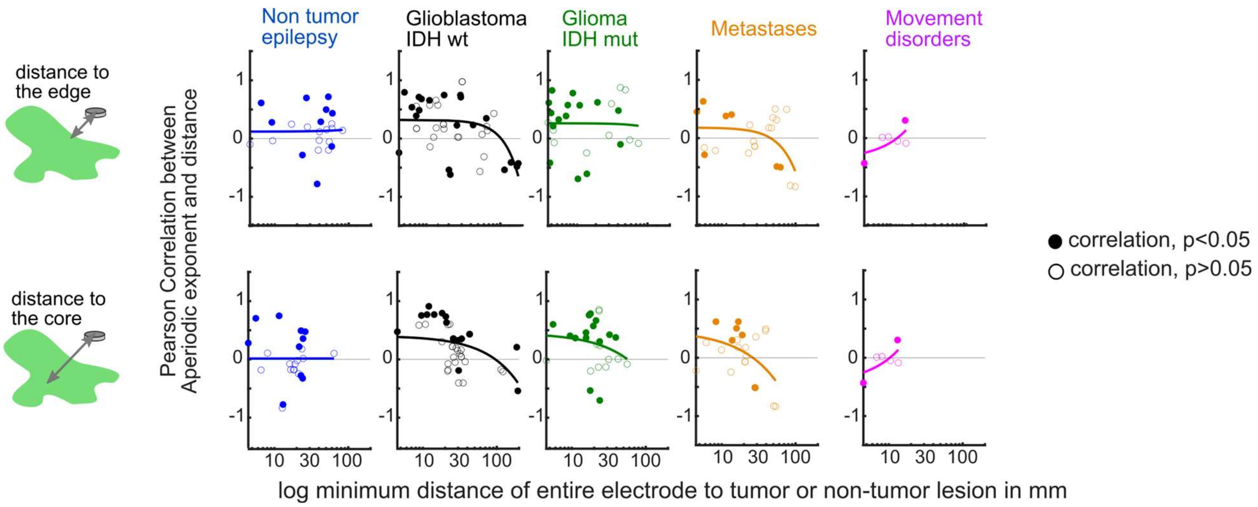

**Supplemental Figure 16. Strength of the effect of distance on the correlation between distance and aperiodic exponent.** Correlation values between aperiodic exponent and distance to tumor edge (top) or core (bottom) versus the log minimum distance of each electrode device to the tumor or non-tumor lesion in mm. Filled in points are correlation values which are significant (Pearson's).

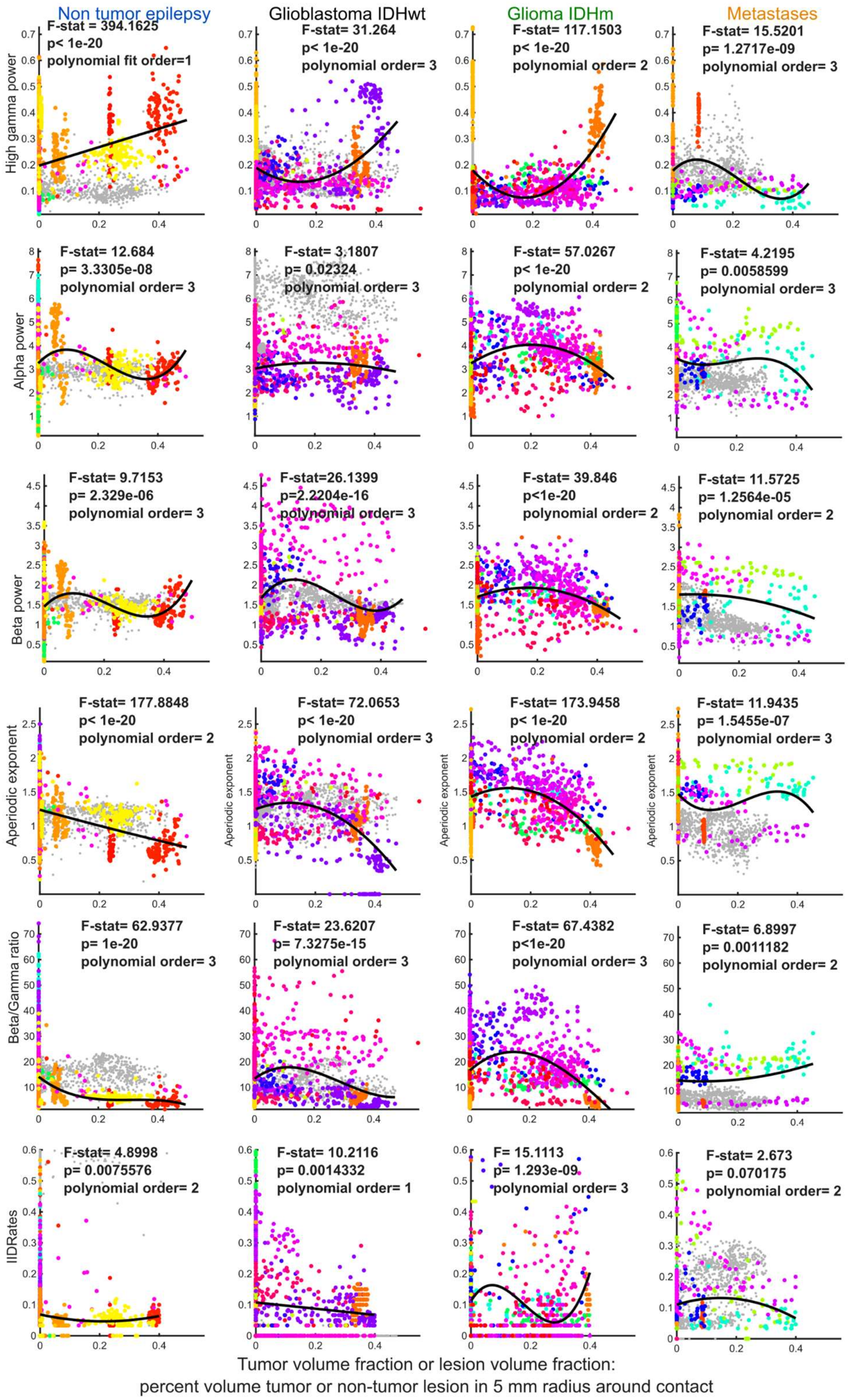

**Supplemental Figure 17. Correlation values between percent volume around electrode contacts intersecting with pathology (tumor or non-tumor lesion) and neurophysiological measures.** F-stat- F-statistic. Lines are polynomial fits with polynomial order indicated in each figure. The multicolored (not grey) dots each indicate a different participant. They grey dots are different participants but include electrodes with >800 contacts. They were not used in the polynomial fit to compensate for any weighting of the fit.

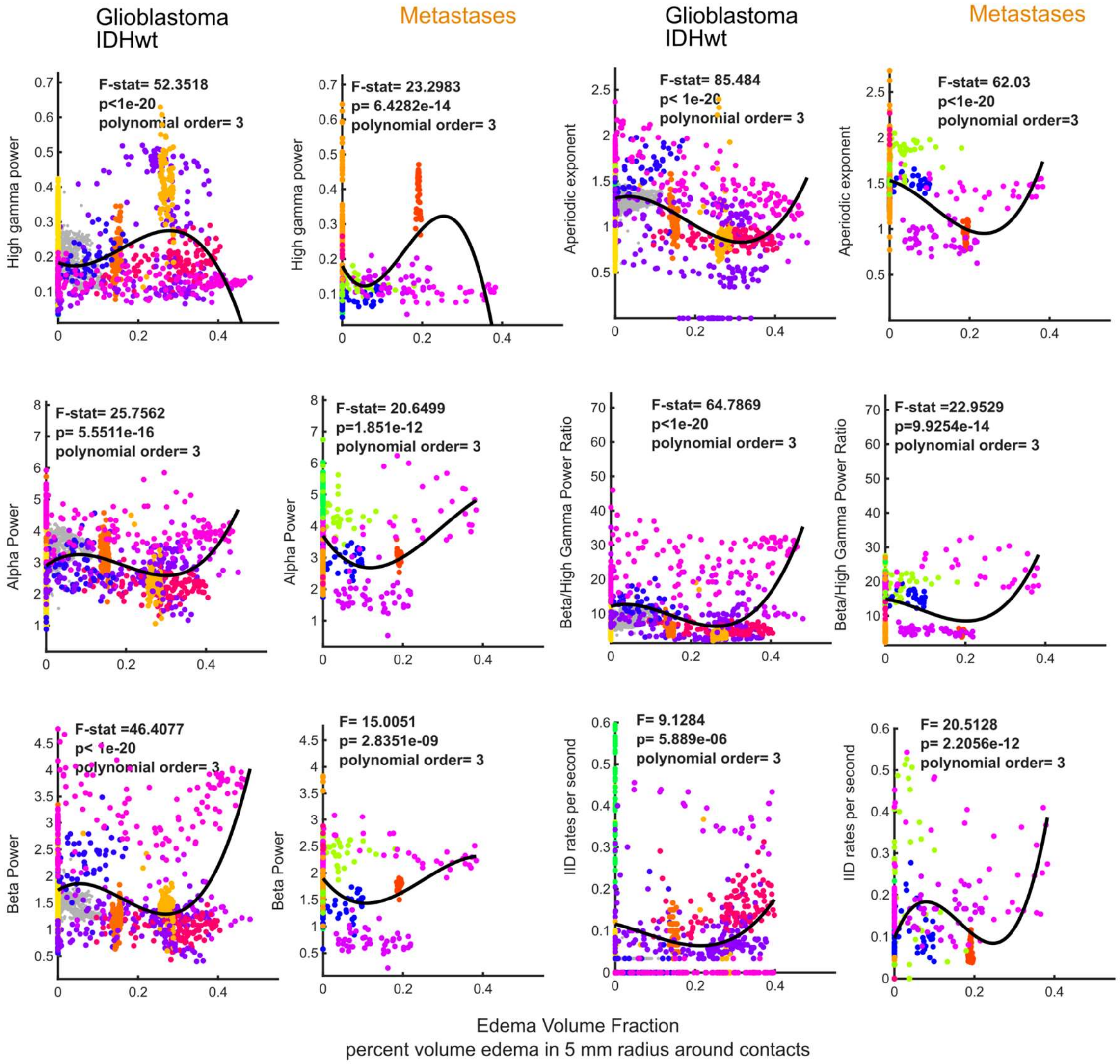

**Supplemental Figure 18. Correlation values between percent volume around electrode contacts intersecting with edema and neurophysiological measures.** F-stat- F-statistic. Lines are polynomial fits with polynomial order indicated in each figure. The multicolored (not grey) dots each indicate a different participant. They grey dots are different participants but include electrodes with >800 contacts. They were not used in the polynomial fit to compensate for any weighting of the fit.

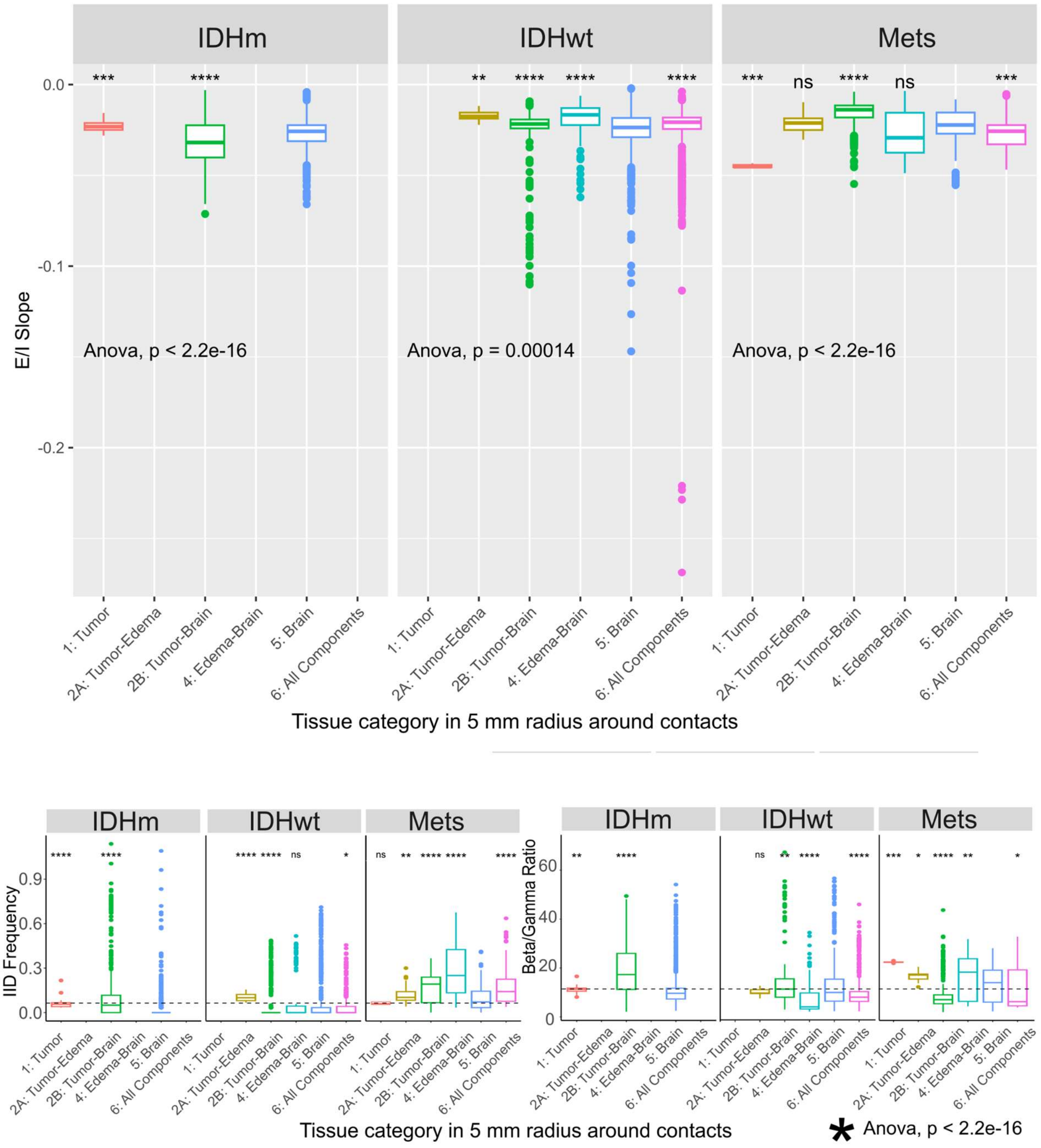

**Supplemental Figure 19. Tissue category in volume around electrode contacts versus excitatory/inhibitory slope (E/I Slope).**

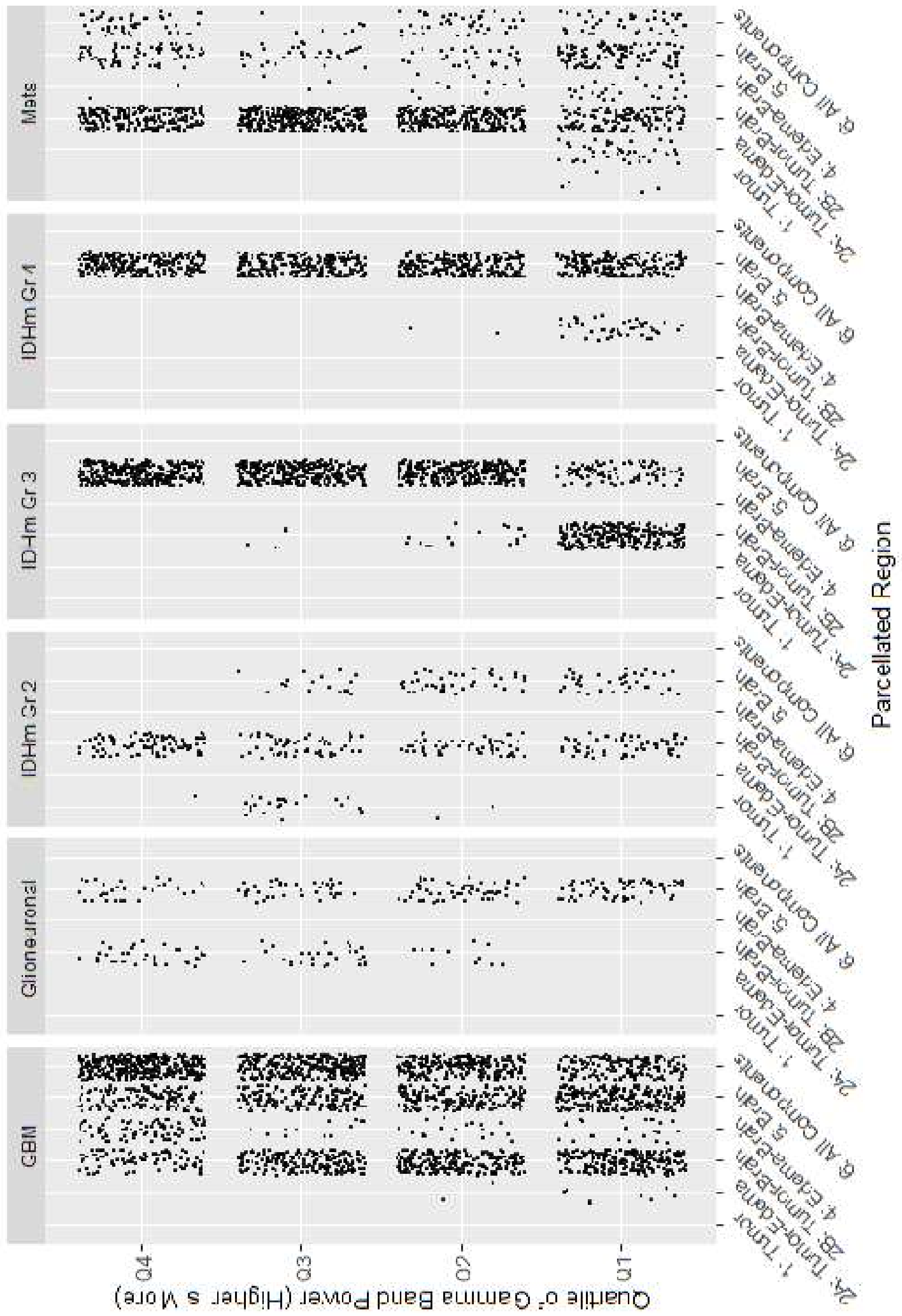

**Supplemental Figure 20. Tissue category in volume around electrode contacts versus high gamma power.**

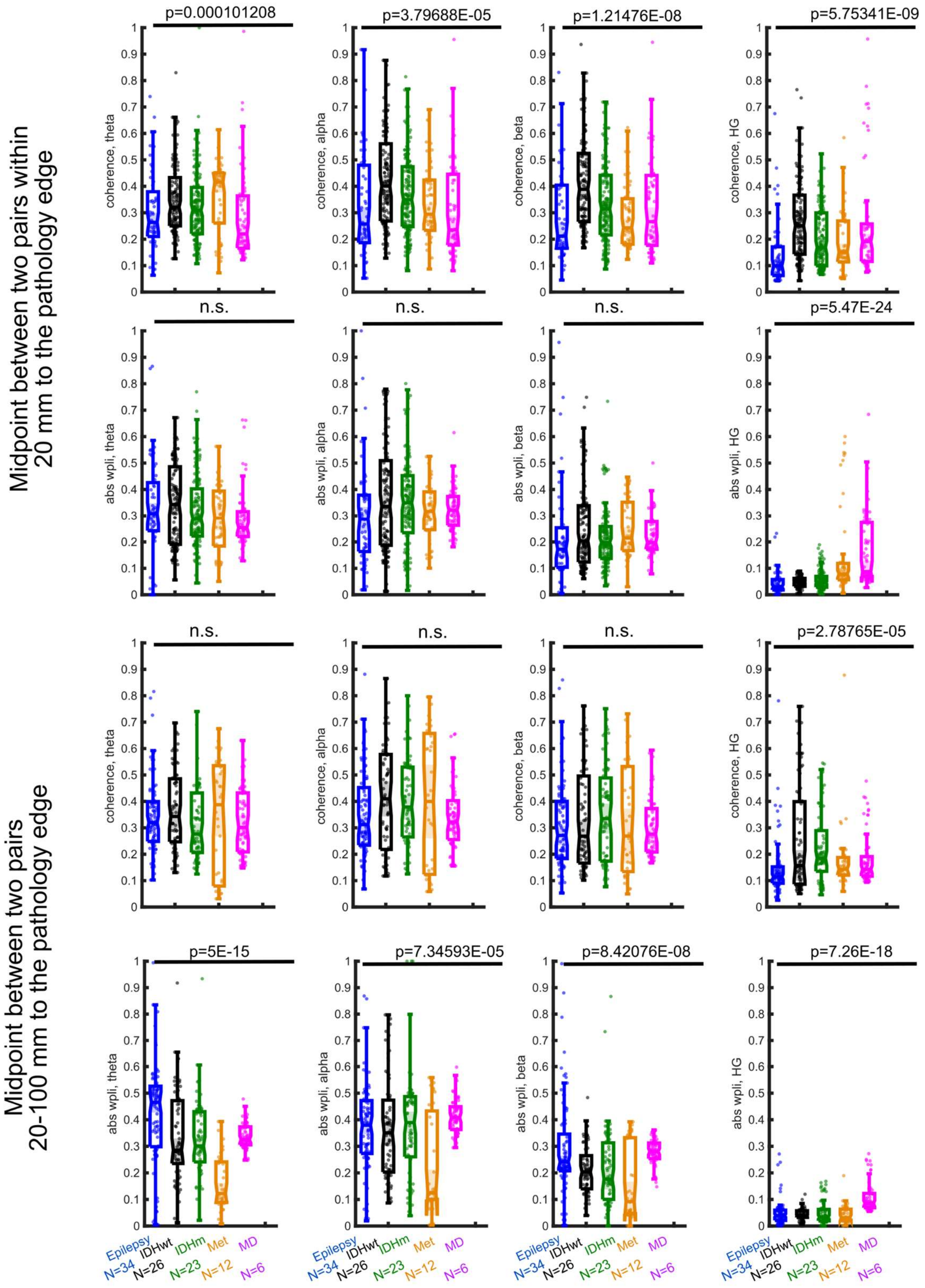

**Supplemental Figure 21. Connectivity differences between contacts at different locations relative to the edge of the pathology.** p-values are for significant differences across groups, Kruskal-Wallis multiple comparisons test. n.s. indicates p> 0.0042.

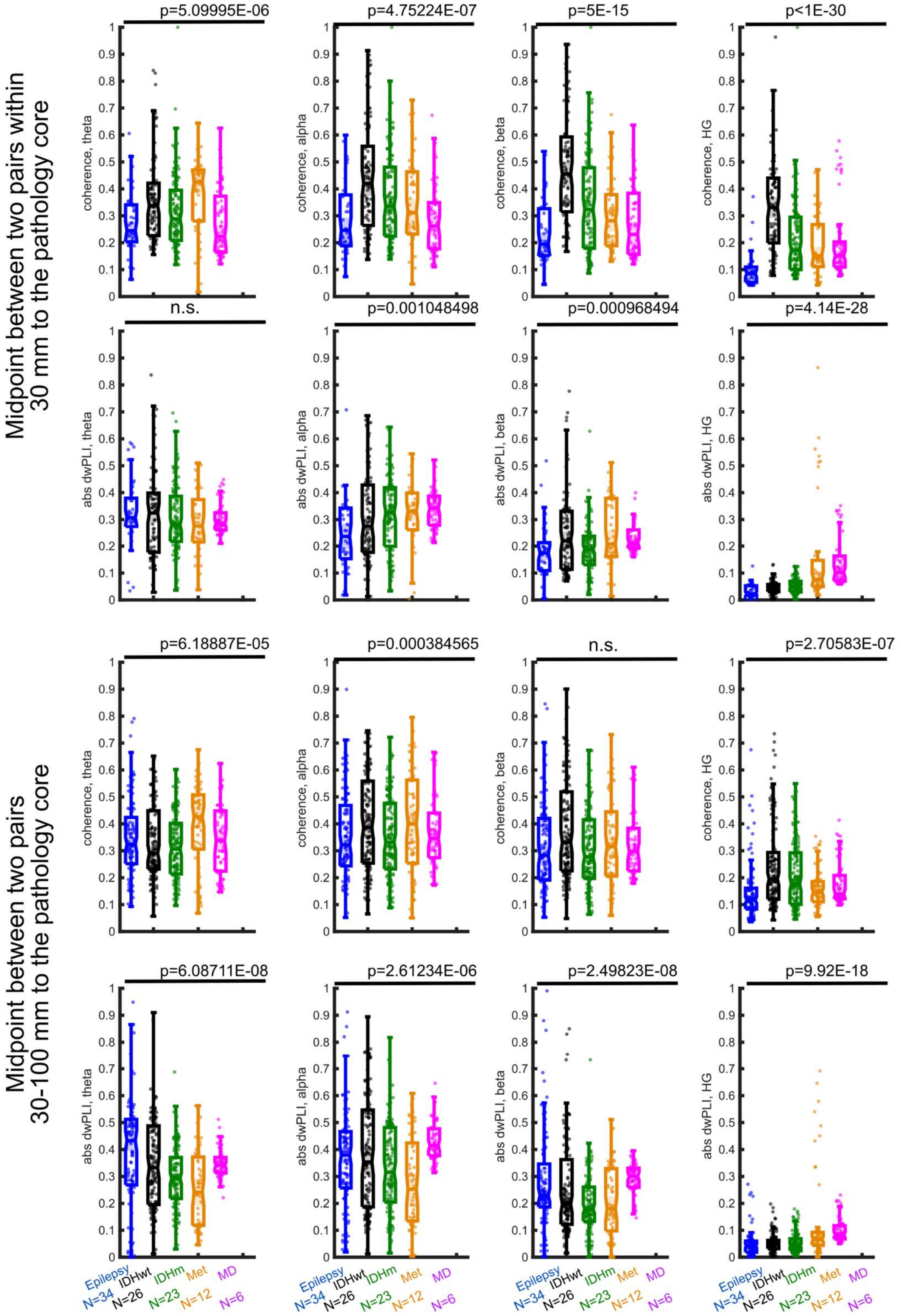

**Supplemental Figure 22. Connectivity differences between contacts at different locations relative to the core of the pathology.** p-values are for significant differences across groups, Kruskal-Wallis test. n.s. indicates p> 0.0042.

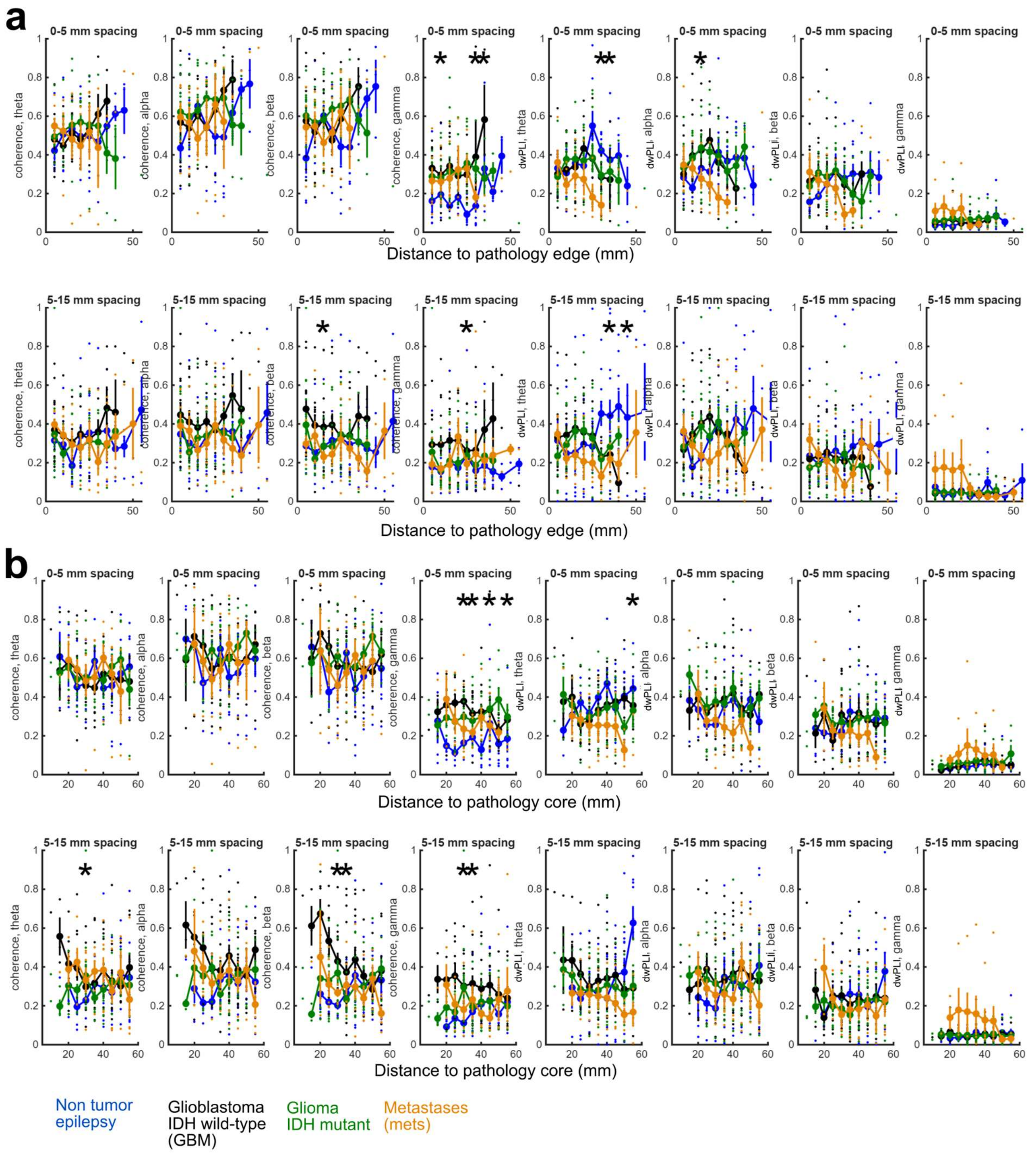

**Supplemental Figure 23. Connectivity differences between contacts at different locations relative to the edge and core of the pathology.** *- p<0.05 significantly different between groups at each binned step and where at least N>5 individuals contributed to each point. Kruskal-Wallis test between diagnoses. Error bars are standard error. Scatter plots are from binned values from individual participants.

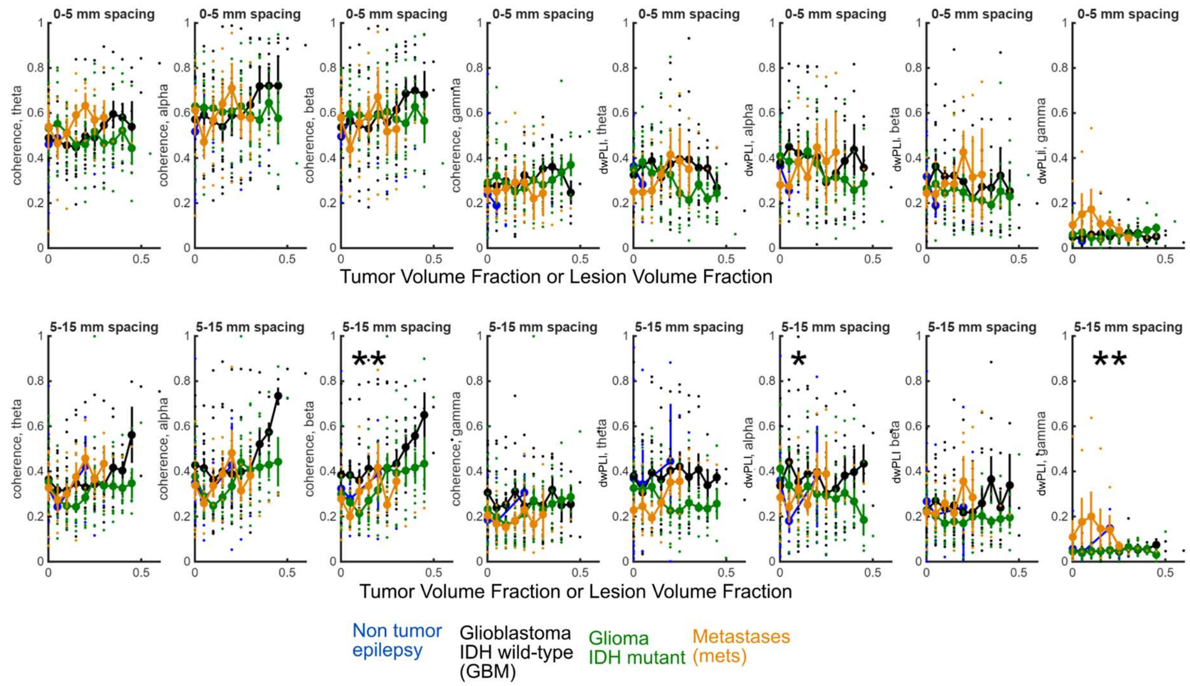

**Supplemental Figure 24. Connectivity differences between contacts at different locations relative to the TVF or LVF.** *- p<0.05 significantly different between groups at each binned step and where at least N>5 individuals contributed to each point. Kruskal-Wallis test between diagnoses. Scatter plots are from binned values from individual participants.

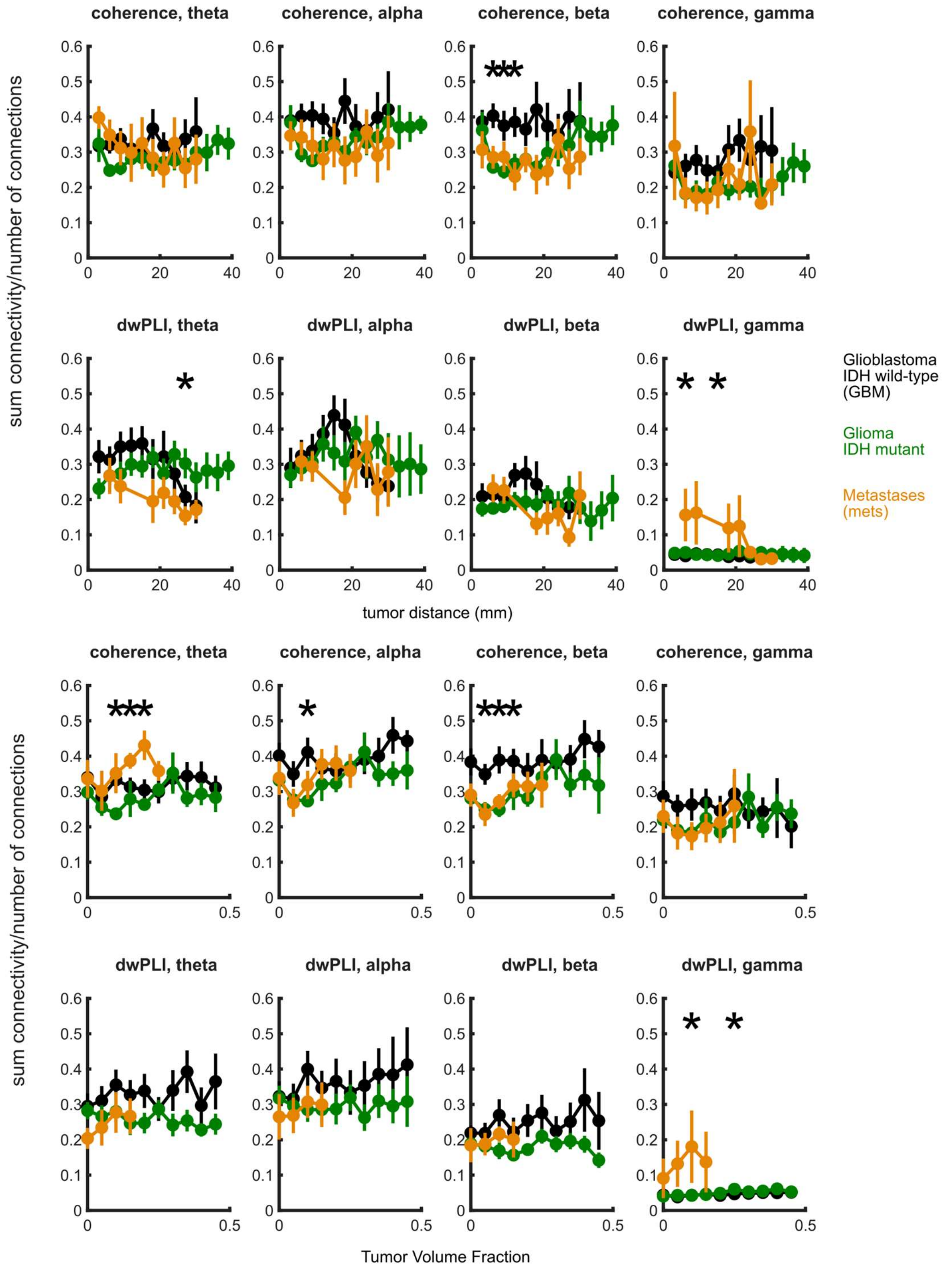

**Supplemental Figure 25. Nodal connectivity differences between contacts at different locations relative to the distance to the tumor and TVF or LVF.** *- $p<0.05$ significantly different between groups at each binned step and where at least N>5 individuals contributed to each point. Kruskal-Wallis test between diagnoses per bin.
